# Population-scale molecular reconstruction of human circadian phase from blood biomarkers

**DOI:** 10.64898/2026.07.08.26356418

**Authors:** Clara Albiñana, Rebecca Richmond, Baihan Wang, Lea Urpa, Jacob J. Crouse, Yanni Zeng, Daniel B. Rosoff, Safia Abdi, FinnGen, Liming Li, Zhengming Chen, Iona Y. Millwood, Hanna M Ollila, Ian B. Hickie, Frédéric Gachon, Achim Kramer, David W. Ray, Naomi R. Wray

## Abstract

Circadian timing influences human physiology and disease risk, yet scalable measures of molecular circadian phase are lacking. Here we infer circadian phase from circulating blood biomarkers in UK Biobank. Among 3,228 plasma biomarkers, 58% exhibit significant diurnal variation, with harmonic modeling identifying acrophase clustering consistent with canonical circadian patterns and independent constant-routine datasets. Machine-learning models trained on plasma proteomics predict sampling time (R²≈0.68) and retain substantial accuracy with ∼60 proteins. We define a novel construct, circadian acceleration (CA), as deviation from the population-average phase; CA is temporally stable, associates with chronotype and shift work, and responds to environmental perturbation. CA is heritable (h²_SNP_≈0.10) and genetically correlated with chronotype and accelerometry-derived sleep traits. These results establish plasma proteomics as a scalable approach for population-level molecular circadian phenotyping.

## Introduction

Temporal changes across the 24-hour day profoundly influence human physiology and behavior (*1*). The circadian clock coordinates gene expression, endocrine secretion, metabolism, immune function, and neural activity across tissues to align with environmental cycles (*2–4*). When internal biological time becomes misaligned with behavioural or external schedules, the risk for cardiometabolic, neuropsychiatric, and inflammatory disorders increases (*5–10*), establishing circadian misalignment as a fundamental determinant of human health.

Despite its importance, a scalable measurement of individual circadian phase remains a major barrier to population-level circadian medicine (*11–14*). The current gold-standard marker, dim-light melatonin onset (DLMO), requires repeated sampling under tightly controlled laboratory conditions, restricting its use to small experimental studies. As a result, large-scale epidemiological studies rely on behavioral proxies such as self-reported chronotype or actigraphy-derived activity timing (*15–18*), which may not fully capture systemic molecular phase.

To address this limitation, molecular approaches have been proposed to infer circadian phase from single samples. Transcriptomics-based methods use rhythmic expression in canonical clock genes or data-driven transcript sets (*14*, *19–23*), while metabolomics-based “molecular timetable” approaches estimate internal time from circulating small molecules (*24*). These approaches demonstrate that the circadian rhythm leaves measurable signatures in peripheral samples, but also highlight that different molecular layers capture distinct aspects of circadian organisation. Transcript levels do not necessarily reflect downstream protein abundance or secretion dynamics, and metabolites can be strongly influenced by short-term behavioural factors such as feeding and activity (*25*, *26*). Despite this, proteomic-based circadian phase predictors remain relatively underexplored, even though circulating proteins are more directly linked to physiological function and integrate signals across tissues.

Large population biobanks provide a unique opportunity to compare circulating molecular markers systematically. Although sampling is cross-sectional at the individual level, dense sampling across clock time in hundreds of thousands of individuals may be sufficient to recover population-level circadian structure and identify which biomarker classes are most informative for phase inference.

Here, we leverage up to 493,493 participants in the UK Biobank (UKB) and external validation cohorts to quantify time-of-day variation across thousands of circulating biomarkers, compare their utility for circadian phase prediction, and define a population-relative measure of circadian misalignment, circadian acceleration (CA). We then examine its temporal stability, environmental sensitivity, and genetic architecture. Together, these analyses establish plasma proteomics as a scalable approach to molecular circadian phenotyping in cohorts far larger than those feasible under laboratory protocols.

## Results

### Overview of the study

We developed a scalable framework to reconstruct molecular circadian phase and quantify circadian misalignment at population scale (Fig. 1). Leveraging up to 493,493 UKB participants (aged 40-70) sampled across an 11-hour daytime window (with 5,000–50,000 samples per clock hour) we first quantified time-of-day variation across 3,228 circulating biomarkers. Despite each participant contributing only a single timepoint per visit, the density of sampling enabled robust harmonic reconstruction of rhythmic structure, which was concordant with laboratory-based independent datasets used for validation. We then trained linear and tree-based supervised machine-learning models to infer molecular circadian phase from plasma profiles and defined circadian acceleration (CA) as deviation from the population-average phase at a given clock time. CA was subsequently evaluated for temporal stability, behavioural and environmental correlates, and genetic architecture.

**Fig. 1.**
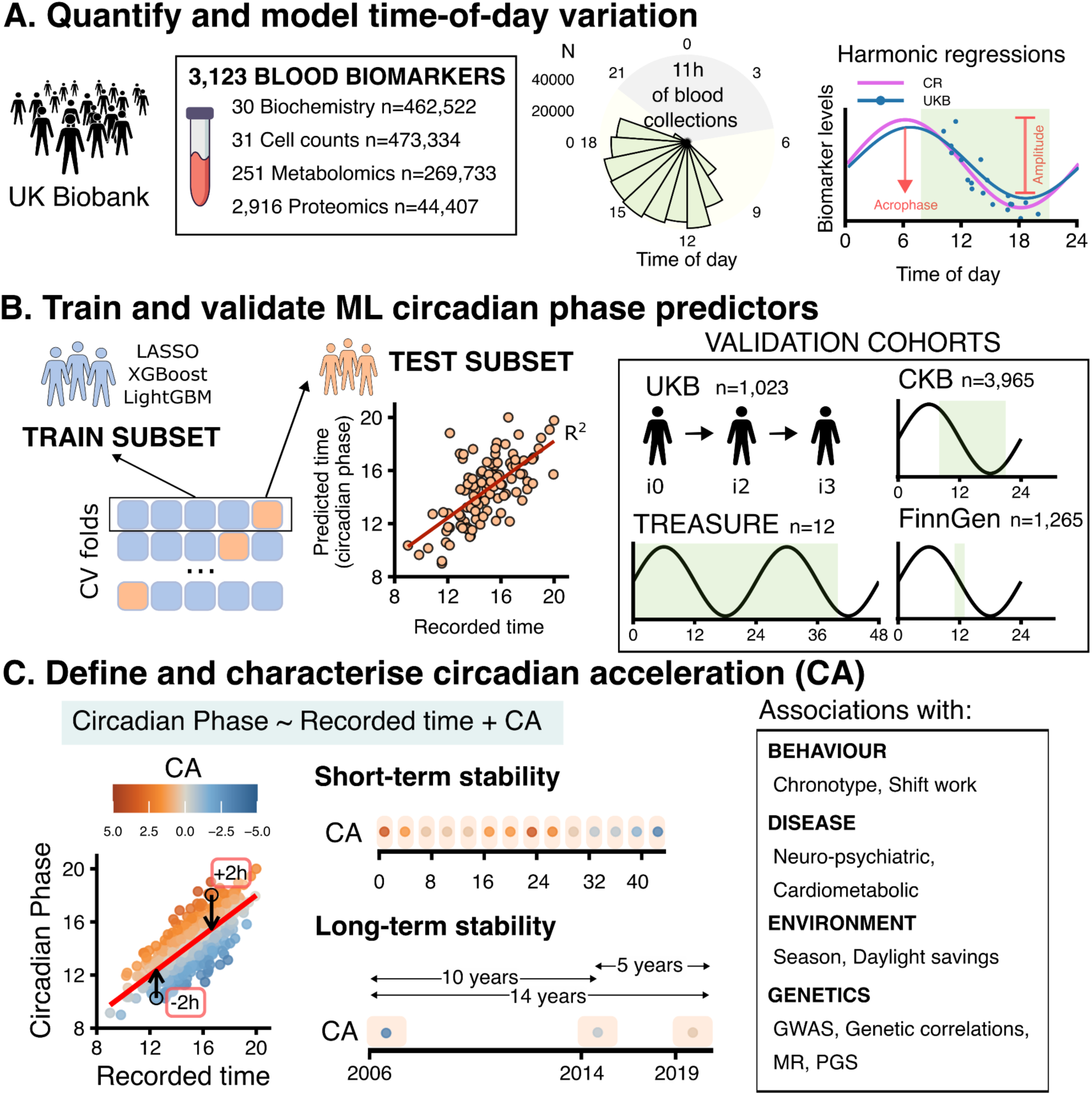
Overview of study design. **A.** Population-scale reconstruction of circadian rhythmicity. We quantified time-of-day variation across 3,228 circulating plasma biomarkers in UK Biobank (UKB), including clinical biochemistry, cell counts, metabolomics, and proteomics. Biomarkers measured at the initial assessment visit (2006–2010) were analysed using recorded sampling time (circular histogram in figure), and harmonic regression was applied to derive acrophase and amplitude parameters. Sample and measurement characteristics are summarised in Table S2. CR: constant routine protocol. The green rectangle indicates blood collection times. **B.** Proteomic circadian phase prediction and validation. We trained and compared multiple machine-learning (ML) algorithms (LASSO, quadratic LASSO, XGBoost, LightGBM) using 5-fold cross-validation to infer molecular circadian phase from plasma biomarkers. Predictors were validated internally using repeated UKB blood samples (repeat assessment, imaging visit, repeat imaging visit) and externally in independent cohorts (China Kadoorie Biobank; CKB, FinnGen, and TREASURE), enabling evaluation across study designs and ancestries. **C.** Circadian acceleration (CA). We derived circadian acceleration (CA) as a population-relative measure of molecular phase misalignment, defined as deviation of predicted circadian phase from the population-average phase at a given clock time. The short and long-term stability of CA was characterised in TREASURE and UKB repeated measures, as well as its phenotypic and genetic associations. GWAS: genome-wide association study; MR: Mendelian randomization; PGS: polygenic score.

### Population-scale time-of-day variation across circulating biomarkers

To quantify rhythmicity in circulating biomarkers, we analyzed 3,228 plasma biomarkers measured in up to 493,493 UKB participants sampled between 09:00 and 20:00 (Table S1). These included 30 clinical biochemistry assays (n=462,522), 31 hematological measures (n=473,334), 251 NMR metabolomic measures (n=269,733), and 2,916 plasma proteins quantified using Olink Explore 3072 (n=44,407).

Time-of-day was significantly associated with 1,857 of 3,228 biomarkers (58%) after false discovery rate (FDR) correction (Data S1). For comparison, sex was significantly associated with 73% of biomarkers, age with 70%, BMI with 83%, smoking status with 60%, and month of assessment with 50%. Time-of-day explained >1% of variance in 134 biomarkers and >5% in 11 biomarkers (Fig. S1).

These strong diurnal signals were observed in immune cell counts (leukocytes, lymphocytes, monocytes, neutrophils, basophils), standard biochemistry measures (phosphate, bilirubin), and numerous proteomic and metabolomic features spanning inflammatory mediators, endocrine regulators, lipid transport proteins, and energy metabolism pathways. These findings confirm pervasive circadian organization across systemic physiology.

### Harmonic modeling reveals coherent phase structure

Hourly means for biomarkers with R2_time-of-day_ >1% (n=134) showed temporal structure consistent with a partial 24-hour oscillation, corresponding to approximately half of the circadian cycle (Fig. S2). We applied first-order harmonic regression to covariate-adjusted biomarker residuals assuming a 24 h period (Data S2). A total of 1,708 biomarkers exhibited significant oscillatory patterns (5% FDR). This included 98% of metabolites, 90% of biochemistry measures, 90% of hematological traits, and 48% of proteins.

Estimated acrophases for all biomarker types clustered around two canonical peaks or “rush hours”: early morning (∼07:00) and early evening (∼19:00), similar to the phase distribution observed for rhythmic proteins in studies with time-resolved sampling (*27*, *28*) (Fig. 2A).

**Fig. 2.**
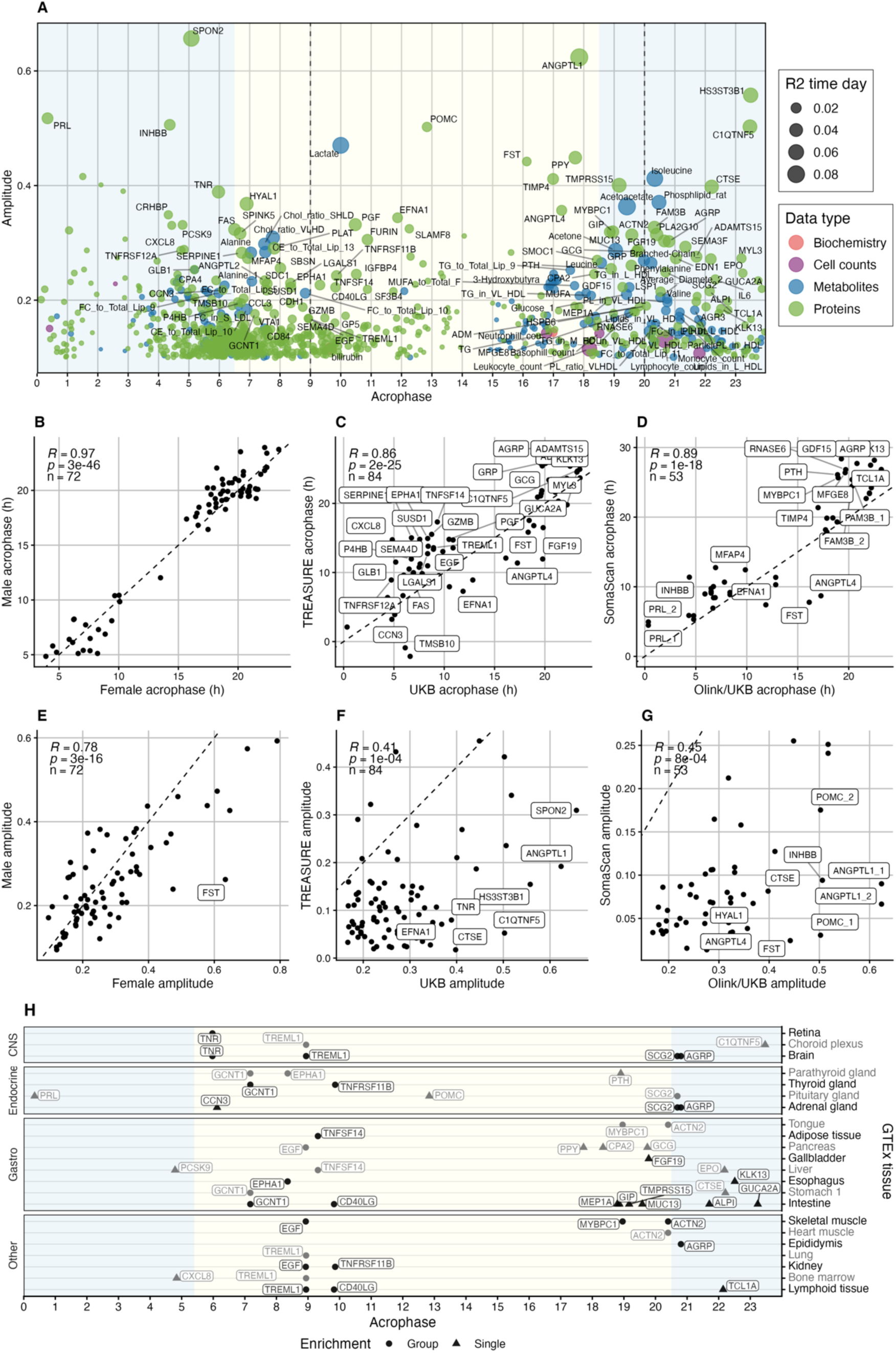
Estimated harmonic parameters for blood biomarkers. **A.** x-axis: The acrophase (time of highest value across 24h period) estimates and y-axis: amplitude (absolute difference between highest and lowest values) estimates for 1,708 significant biomarkers (5% FDR). All results are summarised in Data S2. Labels highlight the subset of 134 biomarkers with R2_time-of-day_ >1%. Dot size indicates R2_time-of-day_ and color indicates biomarker type. Vertical dashed lines at 9:00 and 20:00 indicate UKB blood collection range. Light/Dark shading indicate average sunlight hours in the UK, with yearly sunrise ∼6:30 and sunset ∼18:30. Estimated acrophases for all biomarker types clustered around two canonical peaks or “rush hours”: early morning (∼07:00) and early evening (∼19:00) **B-G.** Biomarker harmonic parameters correlations between sexes (Panels B&E) and independent data using different protein quantification platforms (Panels C&F) and (Panels D&G). The dashed line shows the diagonal. Labels indicate estimate difference >4h (acrophase) and 0.3 (amplitude). Proteins PLAT and GIP missing in TREASURE. **H.** Inferred tissue-of-origin (y-axis) of proteins with 4-fold enrichments of gene expression in GTEx. Note: some proteins are expressed in multiple tissues (1 tissue in ‘Single’, 2-5 tissues in ‘Group’). CNS: central nervous system.

Proteins and metabolites exhibited larger diurnal amplitudes (mean amplitude 0.13–0.14 standard deviation units (SDU)) compared with biochemistry measures and cell counts (0.03–0.05 SDU). Amplitude correlated strongly with variance explained by time-of-day, consistent with larger amplitude oscillations driving greater detectable time-of-day variance (Fig. S3). Estimated amplitudes for some high-sample-size biomarkers (e.g., leukocyte counts) were modest yet explained substantial time-of-day variance, consistent with high statistical power.

### Sex differences in circadian amplitude but not phase

Given known sex differences in circadian biology (*29*), we estimated sex × time interactions among biomarkers with robust rhythmicity (n=131, R²>1%, amplitude>0.1). Significant sex × time interactions were found for 72 biomarkers (Fig. S4).

Acrophase estimates were highly concordant between males and females in these 72 biomarkers (r=0.97, p-value=3×10^−46^; Fig. 2B), indicating conserved peak timing across sexes. In contrast, amplitudes were more moderately correlated (r=0.78, p-value=3×10^−16^; Fig. 2E), with larger oscillation amplitudes in females. These findings are consistent with prior reports of greater melatonin amplitude in females (*30*) and suggest that sex differences in circadian organization may primarily manifest as amplitude modulation rather than phase displacement.

### External validation of protein acrophases

To validate population-derived phase estimates (noting that many were outside the blood sampling time range in our UKB training dataset), we compared acrophases with independent longitudinal proteomic datasets quantified with different proteomic technologies: TREASURE (Olink Explore HT) and Specht *et al*.(*28*)(SomaScan). This analysis was restricted to 86 proteins out of 131 rhythmic biomarkers, as comparable longitudinal datasets for metabolomic, cell counts and biochemistry, measured on standardized platforms, are currently lacking.

In the TREASURE constant-routine cohort (12 participants; 40-hour serum sampling at 3-hour intervals), acrophase estimates for rhythmic proteins were strongly correlated with UKB estimates (Fig. 2C; r=0.86; p-value=2×10^−25^). Similar concordance was observed in the SomaScan-based constant-routine cohort (17 participants; 36-hour plasma sampling at 1-hour intervals), with high correlations across 53 overlapping proteins (Fig. 2D; r=0.89; p-value=1×10^−18^), supporting consistent phase structure across platforms.

Amplitude estimates were also significantly correlated in both datasets (TREASURE: Fig. 2F; r=0.41, p-value=1×10^−4^; SomaScan: Fig. 2G; r=0.45, p-value=8×10^−4^), with systematically larger amplitudes observed in UKB.

Despite UKB sampling being restricted to daytime hours, 75% of inferred acrophases fell outside the 09:00–20:00 window, indicating that daytime cross-sectional data captures sufficient information to reconstruct full circadian phase structure. The strong concordance of both phase and amplitude estimates in independent constant-routine datasets further supports the validity of this approach, demonstrating that population-scale cross-sectional modeling can robustly recover biologically meaningful circadian organization.

### Plasma proteome captures multi-organ circadian organization

To investigate the tissue origins of rhythmic plasma signals, we mapped proteins to GTEx transcript enrichment profiles across 54 tissues. Of 1,406 rhythmic proteins, 746 were classified as tissue-enriched and spanned 34 tissues, underscoring the multi-organ contribution to circulating circadian rhythms. On average, 42% of enriched proteins per tissue showed significant rhythmicity, with prominent representation from brain, liver, and lymphoid tissues (Fig. S5).

Focusing on the 86 proteins with the strongest rhythmicity (high amplitude and variance explained), we observed broad representation across 22 tissues (Fig. 2H), reinforcing the multi-organ nature of circulating rhythms. Among these, pituitary-associated proteins such as prolactin (PRL) and proopiomelanocortin (POMC) showed distinct acrophases consistent with known dynamics of the hypothalamic pituitary adrenal axis.

More broadly, rhythmic proteins were distributed across metabolic (liver, pancreas, intestine) and immune-related tissues, pointing to coordinated circadian organisation across endocrine, metabolic, and inflammatory pathways. Together, these results support plasma proteomics as a systems-level readout of circadian timing, integrating signals from distributed peripheral clocks.

### Proteomic prediction of circadian phase

We next developed machine-learning models to predict sampling time from biomarker profiles. Four algorithms (LASSO, quadratic LASSO, XGBoost, LightGBM) were trained using 5-fold cross-validation. As all approaches yielded similar performance, we averaged their predictions.

Proteomics achieved the highest mean predictive performance (R²=0.68), substantially outperforming metabolomics (R²=0.42), biochemistry (R²=0.17), and cell counts (R²=0.15). Modeling all biomarkers jointly increased performance modestly (multi-platform: R²=0.71) (Fig. 3A, Fig. S6-8, Supplementary Text S2).

**Fig 3.**
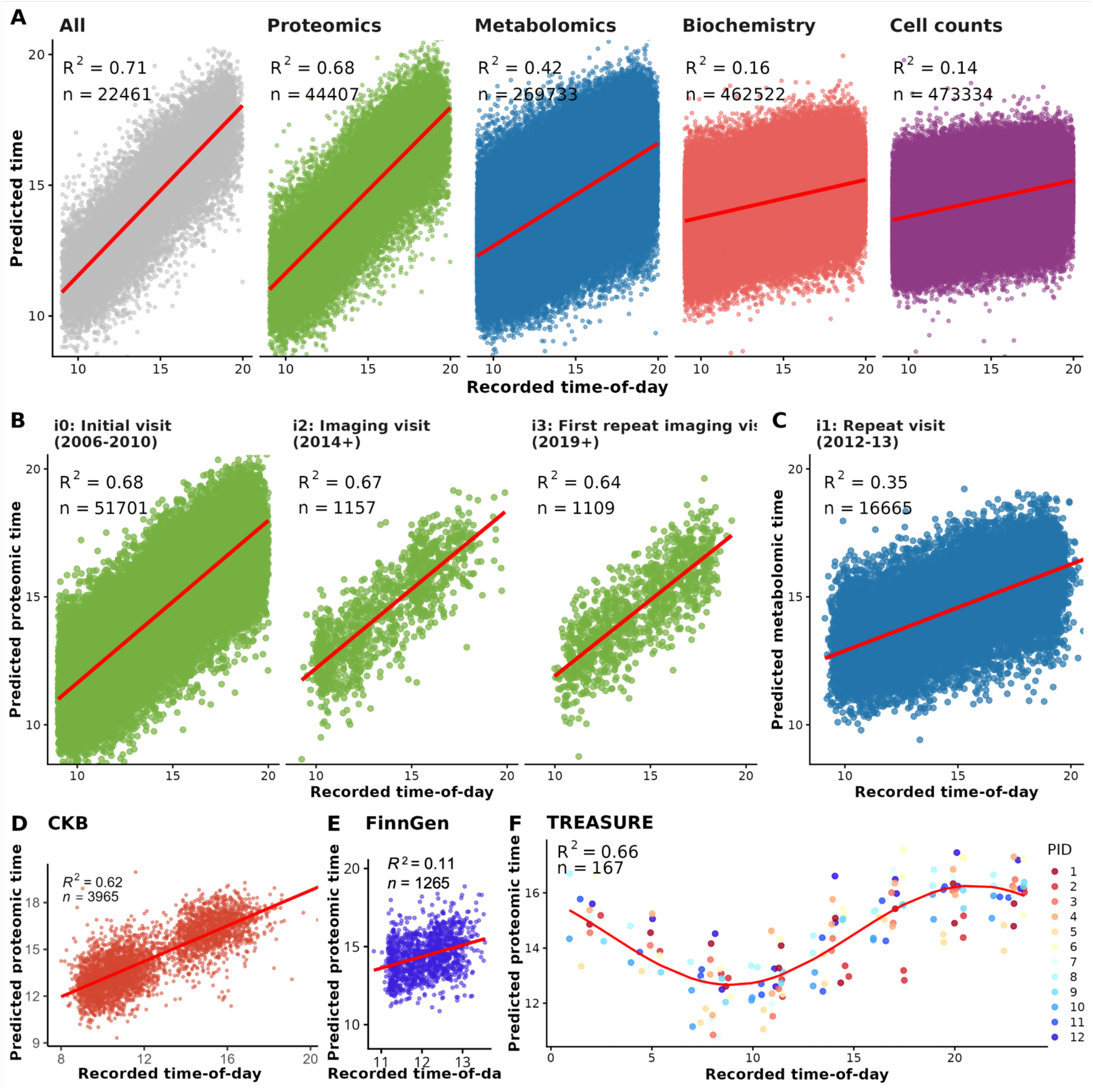
Prediction of blood sampling time-of-day from blood biomarkers. Recorded time-of-day of blood sampling (x axis) and blood biomarker predicted time-of-day (y axis). All predicted times shown are the mean of the four prediction models: LASSO, quadratic LASSO, XGBoost and LightGBM. The red line shows the linear regression line fitted with all data points (cosinor model in 4D). **A.** UKB time prediction comparison between biomarker data types, trained using 5-fold cross validation in the range 9h to 20h. The data in the figure shows all 5 cross-validation subsets. **B.** Prediction validation on UKB repeated proteomics. Time-of-day predictions for individuals with repeated proteomic measures in instance 2 (i2) and instance 3 (i3). The repeated measures of Olink proteomics only included half the panels from the initial assessment (∼1,500 proteins instead of ∼3,000). We retrained the four algorithms on instance 0 (i0) data to only include proteins with repeated measures (See Methods). These proteins were available for a larger subset of the i0 individuals (n=51,701). **C.** Prediction validation on UKB repeated metabolomics. Time-of-day predictions for individuals with repeated metabolomic measures in instance 1 (i1). Proteomic external validation using **D.** CKB, **E.** FinnGen and **F.** TREASURE. Recorded time included samples collected from participant ID (PID) on Days 1, 2 & 3, resulting in up to 3 predictions per time-point and sample.

Prediction accuracy was validated in UKB participants with repeated blood sampling (Fig. S9). After retraining models using the 1,458 proteins shared across visits, proteomic prediction performance remained stable (R² = 0.68 at baseline; R² = 0.67 and 0.64 at repeat visits) (Fig. 3B), while metabolomic prediction showed modest attenuation (R² = 0.42 at baseline; R² = 0.36 at repeat) (Fig. 3C). Prediction accuracy was highest for European-ancestry participants, and decreased modestly for other ancestries (Fig. S10).

Robustness was assessed in three independent cohorts with different study designs (CKB, FinnGen, and TREASURE) and recorded sampling times (Table S2, Fig. S12). Models trained in UKB generalized well (R²=0.62 in CKB; R² = 0.66 in TREASURE), whereas performance in FinnGen was lower (R²=0.11), likely reflecting its narrow sampling window (11:00–14:00) (Fig. 3D-F). Prediction accuracy was also robust to covariate adjustment strategy (no-covariates vs technical covariates vs full adjustment; mean R² 0.68–0.69) (Supplementary Text S3).

To evaluate translational feasibility, we assessed whether accurate phase prediction could be achieved with smaller protein panels. Performance plateaued below ∼1,000 proteins and remained substantial even when restricted to ∼60 features (R²=0.40), indicating that high-dimensional proteomics is not strictly required for meaningful circadian inference (Supplementary Text S4).

### Circadian acceleration: a population-relative biomarker of circadian misalignment

We defined the novel construct of circadian acceleration (CA) as a population-relative measure of molecular phase misalignment, computed as the residual of predicted time regressed on recorded time. CA was calculated in 51,701 UKB participants (mean 0; IQR −48 to +49 minutes).

We assessed the longitudinal consistency of CA using repeated proteomic measurements in the same individuals (Fig. 4A). CA showed moderate within-person stability that declined with increasing follow-up years: Pearson’s r = 0.45 at 3.5 years, and r = 0.35 both at 9.2 and 12.6 years. For comparison, correlations in recorded sampling time within individuals were smaller (r=0.20 at 3.5 years and r=0.10 beyond 9 years, Fig. 4A). While appointment times were partly scheduled by UKB rather than purely random, these lower correlations suggest that CA captures more persistent individual-level differences than variation driven by sampling time alone. This interpretation is supported by a mixed-effects model, which yielded an intraclass correlation coefficient (ICC) of 0.32, indicating that ∼32% of variance in CA reflects stable between-individual differences.

**Fig 4.**
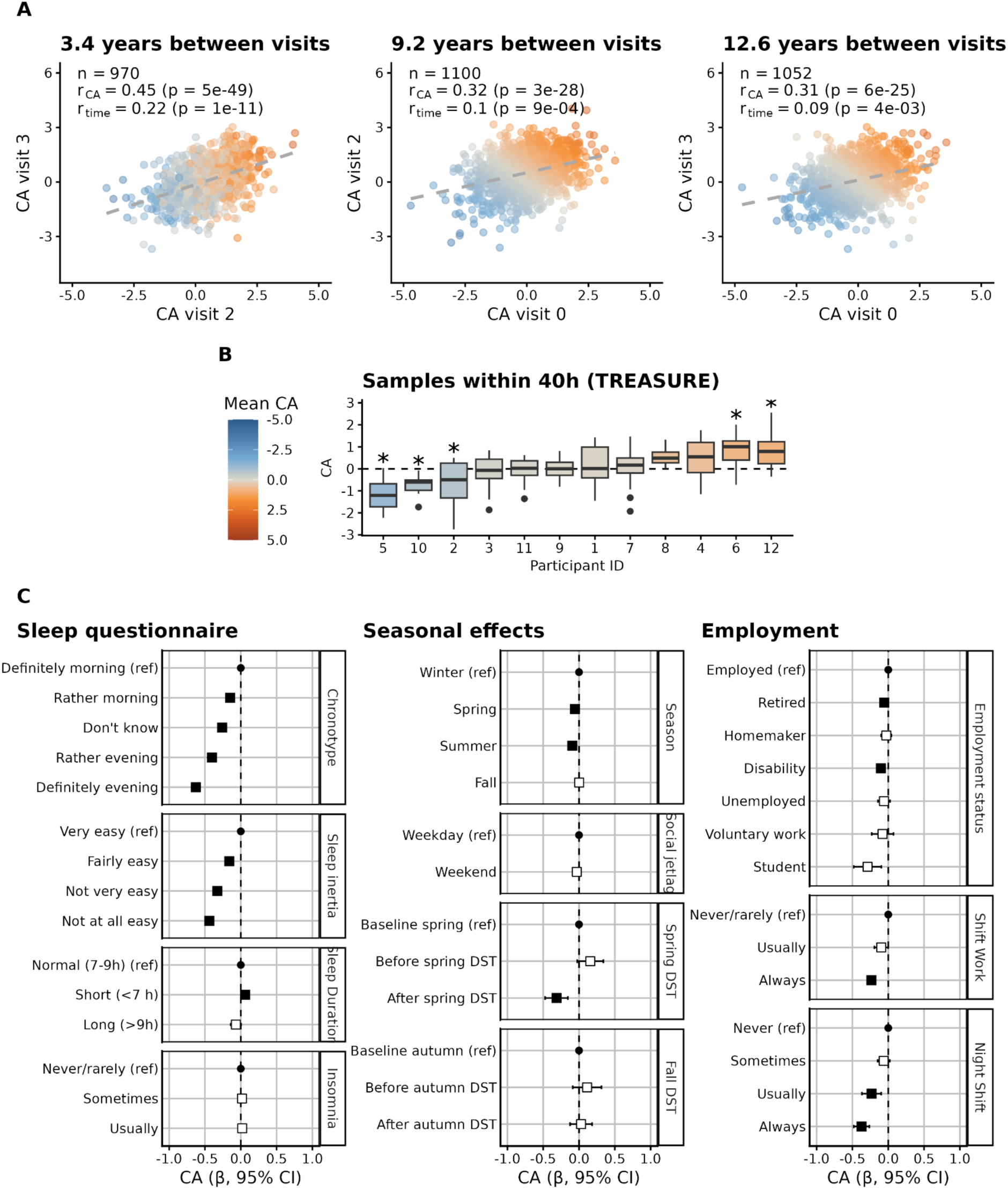
Blood circadian acceleration (CA) stability over time and phenotypic correlates. **A.** Correlation plots between CA estimates across the three UKB timepoints with repeated proteomics data. Pearson’s correlation is shown between individual CA estimates (r_CA_) and visit time of day (r_time_). Visits are compared according to the number of years that passed between them: 3.4 years (SD ±1.6), 9.2 years (SD±1.8) and 12.6 years (SD±1.0) follow-up. **B.** Distribution of individual CA estimates (from UKB predictors) over 40 consecutive hours in TREASURE. Each of the 12 individuals (x-axis) shows a distribution of CA (y-axis) for 14 time points. Individuals with mean CA significantly different from 0 are marked as *, at FDR 5%. **C.** CA phenotypic associations with self-reported measures of sleep, seasonal effects and employment. All CA effect sizes (x-axis) come from linear regression models, adjusted for sex, age, assessment centre and the first 20 genetic principal components. A black square indicates the association was FDR significant. Beta estimates and 95% CI are shown in the x axis in hours.

Short-term stability was evaluated in the TREASURE constant-routine dataset using prediction weights trained in UKB (Fig 4B). CA also showed moderate short-term stability within 40h (ICC = 0.37), with consistent positive or negative CA across repeated measurements for several participants. Together, these analyses indicate that CA captures reproducible inter-individual differences in molecular circadian phase alongside appreciable within-individual variation.

### CA correlates with environmental and behavioral factors

We characterised the relationship between CA and a range of demographic, employment, behavioral, self-report and disease measures (Data S3). We report only selected key results here; full phenotypic and disease association outputs are provided in Supplementary Text S5.

CA showed no association with age, sex or genetic ancestry. In contrast, it was strongly associated with self-reported sleep and circadian traits (Fig. 4C). Evening chronotypes were delayed by 38 min relative to morning types, with a graded shift across intermediate categories. Using the composite rMEQ chronotype score (*31*), defined by combining five sleep-related questions, this difference increased to 81 minutes, indicating a stronger alignment with underlying circadian preference (Fig. S14). Greater sleep inertia (difficulty waking) was also associated with delayed CA (up to 26 min for “not at all easy” versus “very easy”), whereas shorter sleep duration showed modest effects (4 min delay). W No association was observed between CA and self-reported insomnia.

Seasonal and social timing effects were detectable at population scale (Fig. 4C). Relative to winter, CA was delayed in summer (6 min) and spring (3 min), suggesting responsiveness to photoperiod. A pronounced acute effect was observed after the spring daylight saving time transition (1-hour clock advance): participants sampled on the following Monday-Tuesday showed a 19 min delay, while no comparable shift was detected after the autumn transition.

We observed differences in CA across employment and shift-work status (Fig. 4C). Compared with employed participants, retired individuals and those self-reporting disability showed modest delays in CA (3 and 6 minutes, respectively). Among employed participants, shift work was associated with delayed CA, with the largest shifts observed in consistent night workers (22 minutes), compared to those who usually worked nights (14 minutes) or always did shift work overall (14 minutes). Notably, persistent night shift work was associated with a 20-minute delay in CA across chronotypes, such that morning chronotypes working permanent night shifts exhibited levels of misalignment comparable to evening chronotypes who never worked shifts (Fig. S15).

Although modest in magnitude, these effects were detectable across ∼50,000 individuals, demonstrating sensitivity of CA to real-world circadian perturbations.

### Genetic architecture of circadian acceleration

SNP-based heritability of CA was estimated at 0.10 (SE=0.01). Genome-wide association analysis identified 1,825 genome-wide significant variants, resolving to 20 independent signals after conditional analysis (Fig. 5A, Supplementary Text S6.1).

**Fig. 5.**
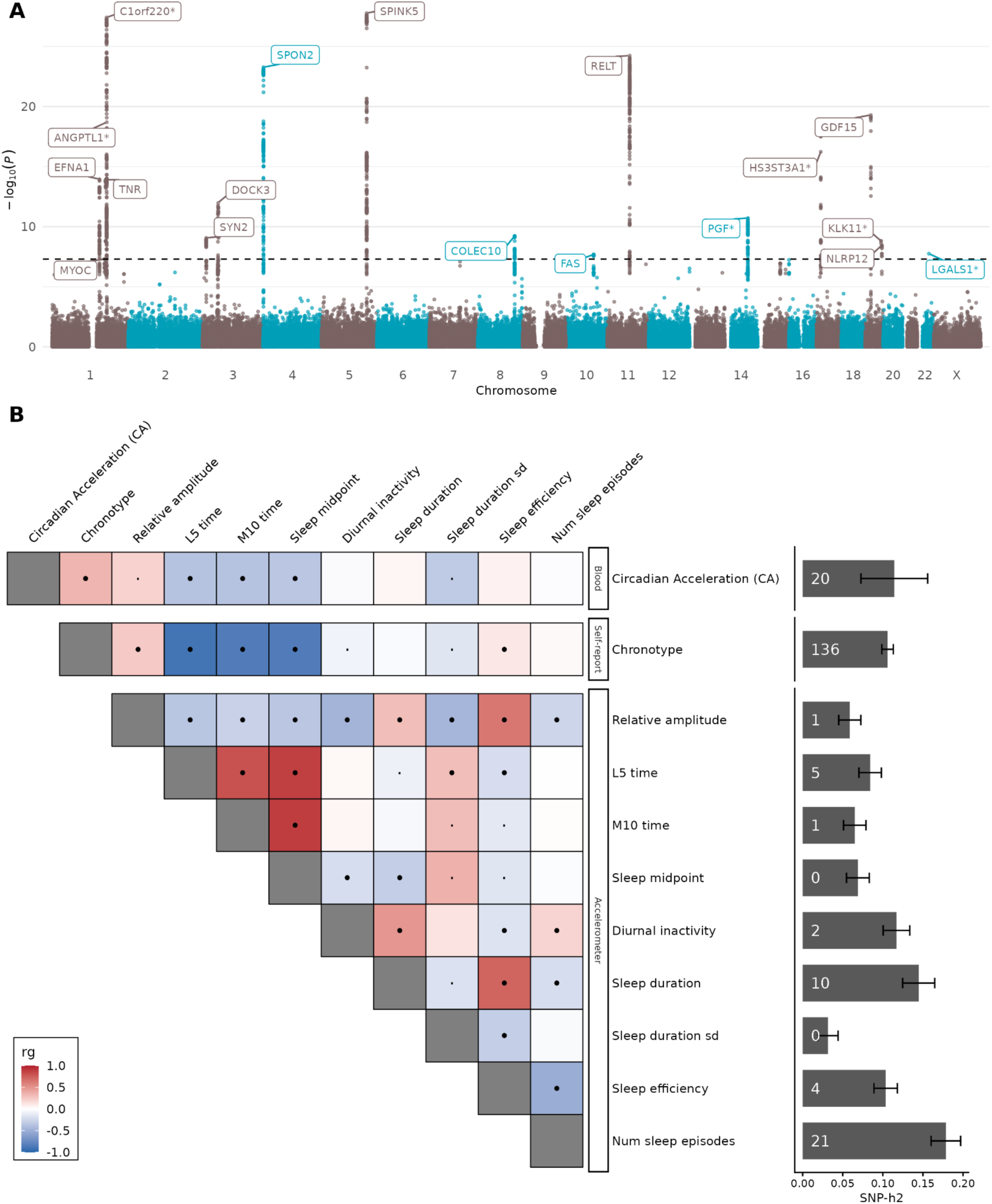
Genetic architecture of blood circadian acceleration (CA). **A.** Manhattan plot for CA GWAS in N=44,529 individuals of European ancestry, highlighting the nearest gene of 20 independent variants across 11 chromosomes. **B.** Genetic correlations between CA and other circadian phase estimates from self-reported and accelerometer-based sources. Dot size indicates significance threshold (·) p-value < 0.05 and (**•**) FDR significant. Bars indicate trait SNP-h2 and number of conditional and joint association analysis (COJO) independent SNPs.

Fifteen of 20 lead variants mapped to genes encoding circulating proteins included in the phase prediction model, indicating enrichment for protein *cis* quantitative trait loci (Supplementary Text S6.2-S6.3). Our results highlight that genetic variants in highly rhythmic proteins can directly impact the protein circadian parameter estimates and, consequently, the individual’s molecular misalignment (CA) values (Fig. S17, Supplementary Text S6.4).

Genetic correlation analyses suggest shared genetic architecture between CA and self-reported chronotype (r_g_=0.34) and accelerometer-derived sleep timing traits (L5 timing r_g_=-0.37, M10 timing r_g_=-0.37 and sleep midpoint r_g_=-0.35) (Fig. 5B). We found no significant genetic correlations between CA and 16 diseases and sleep traits analysed (Supplementary Text S6.5, Data S5-6). Mendelian randomization supported a causal effect of chronotype on CA but not vice versa, providing evidence that chronotype has downstream effects on blood-based biomarkers (Fig. S19, Supplementary Text S6.6). CA had no significant effect on the accelerometry-derived traits (Fig. S20), and we had no robust genetic variants to test the reverse causal relationship.

Finally, CA polygenic scores had high prediction accuracy across the repeated measures (R2=4%, 2% and 3%, respectively for visits 0, 2 and 3), which partially explains the stability of CA over time. We further validated the CA PRS by testing its association with self-reported chronotype in the full UK Biobank sample of European ancestry (n=381,449, excluding proteomics), observing a significant dose–response relationship across chronotype categories (Fig. S19; Supplementary Text S6.6).

## Discussion

We systematically characterised time-of-day variation across 3,228 circulating biomarkers in the UKB and demonstrated that plasma-derived molecular signatures can be used to infer circadian phase at population scale. More than half of the measured circulating biomarkers exhibited significant rhythmic structure, and high-dimensional proteomic data enabled accurate prediction of sampling time (R²=0.68), with high degree of transferability to independent cohorts with different ancestries, ages and study designs.

Circadian research has traditionally relied on tightly controlled laboratory protocols to separate endogenous rhythms from those driven by behaviour or environment (*21*, *28*, *32–35*). Rather than sampling individuals intensively over 24h, we leverage large-scale biobank data in which participants are sampled across a wide range of clock times. While this design cannot reconstruct individual trajectories or fully disentangle behavioural effects from biological rhythms, its unprecedented scale allows for robust estimation of harmonic parameters, which we validated against 24h constant-routine datasets.

Consistent with established circadian biology, we observed clear rhythmic patterns in circulating hormones and broader evidence of coordinated activity across multiple organs, reflecting contributions from both central and peripheral clocks. Because circulating biomarkers integrate endogenous clock signals with behavioural and environmental influences such as feeding and activity (*27*), this framework captures circadian organisation as it operates in real-world physiology, complementing insights from tightly controlled laboratory studies. The integrative nature of the circulating proteome may also explain why proteomics emerged as the most informative platform for circadian phase inference, outperforming clinical biochemistry, blood cell counts, and metabolomics. Notably, predictive performance plateaued at ∼1,000 proteins and remained substantial with ∼60 proteins, indicating that scalable circadian phase estimation may ultimately be feasible using a targeted protein assay. With the forthcoming expansion of the UKB proteomics dataset, including repeated measurements, our approach lays the groundwork for future molecular circadian phenotyping in half a million people.

We introduce circadian acceleration (CA) as a population-relative measure of circadian misalignment. The current gold-standard measures for circadian phase; DLMO (dim-light melatonin onset), is estimated from each individual’s melatonin curve under controlled dim-light conditions. Our CA results strongly aligned with DLMO: CA differed by approximately one hour between evening and morning chronotypes, mirroring the direction of chronotype phase differences reported for DLMO (*36–38*) and for transcriptomic-based predictors of DLMO (*39*). Sleep inertia results were also concordant with DLMO, with delayed CA associated with greater sleep inertia (*40*). Consistent with prior literature (*41*), we observed higher circadian amplitudes in females, but no significant phase differences among the top rhythmic biomarkers. This aligns with reports of greater melatonin amplitude (*30*) and more sustained transcriptomic rhythmicity in females (*29*). We did not find an association between CA and sex. One possible explanation is that sex differences in DLMO have been reported in participants with age <34 years (*30*, *42*, *43*), while age of UKB participants ranges 40-70. It is relevant to note that the DLMO datasets are <50 participants, vs. 50,000 in the UKB, providing much greater power to estimate population-level structure and account for potential confounding. A recent study combined and reanalysed data from 150 DLMO publications on healthy participants, reporting no DLMO differences in sex across age groups (*38*). That study also found extreme DLMO variability within age groups. Overall, the latest DLMO values were observed in adolescence, followed by a gradual advance with age until the 70s. Consistent with this, other work has shown that the largest shift in chronotype occurs between ages 15 and 25 (*44*). We also did not detect a significant association between CA and age, but this may reflect the restricted older age range of UK Biobank participants. Evaluating CA in cohorts that include adolescents and young adults will be important for defining its full age-related trajectory.

CA proved to be a heritable trait, with ∼10% of its variance explained by common genetic variation. Many of the strongest GWAS signals mapped to genes encoding circulating proteins included in the phase prediction model, indicating that genetic regulation of these proteins contributes directly to the CA signal. One plausible mechanism is through baseline protein abundance: pQTLs that increase or decrease circulating protein levels may change the signal-to-noise ratio of rhythmic measurements, making temporal patterns easier or harder to detect. In some cases, genetic effects could also influence the amplitude of rhythmic protein variation itself. This distinction is important because accurate phase inference depends on sufficient rhythmic signal. Individuals with lower-amplitude or lower-abundance proteomic oscillations may therefore have less precise phase estimates. As a result, extreme CA values may reflect not only true phase advance or delay, but in some cases weaker rhythmic signal. Future work, particularly with repeated measurements, should aim to disentangle phase timing from rhythm strength so that these can be modelled as complementary dimensions of circadian misalignment.

Through genetic correlations, we report that blood-based CA captures characteristics of both self-reported chronotype and accelerometer-based measures of sleep and activity, including most-active and least-active periods and sleep timing. This replicability across data domains links the behavioral nature of chronotype with a biological, systemic signal in blood. This link is further strengthened by our Mendelian randomization findings, indicating that chronotype influences CA, but not vice versa. These results deepen our understanding of chronotype differences by adding a multi-omic component beyond melatonin, but more research is needed to determine if the downstream effects on CA are rooted in genetic or environmental differences (e.g., light exposure) across chronotypes.

CA captures both heritable and dynamic state-dependent variation. Our results also show that CA is responsive to environmental perturbations, including night shift work, daylight saving time (DST) transitions and seasonal changes. Individuals who consistently worked night shifts exhibited delayed CA irrespective of baseline chronotype, which is also observed in terms of DLMO (*45*). Interestingly, morning chronotypes continuously working night-shifts showed misalignment levels comparable to evening chronotypes who never worked shifts, suggesting that conventional work schedules are aligned with early chronotype phasing, and explaining the basis of increased disease risk seen in late chronotypes (*5–10*). We also observed a pronounced DST effect: participants whose blood was drawn on the Monday and Tuesday immediately following the spring DST transition (1-hour clock advance) exhibited 20min delay in CA, whereas no comparable shift was observed following the autumn transition (1-hour clock delay). This asymmetry is consistent with evidence that humans generally adapt more readily to phase delays than to phase advances (*46–48*). However, these effects were not explained by reduced sleep alone, as short sleep duration was associated with a delay of 4min in CA compared with those sleeping 7–9 hours. These results suggest that environmental scheduling exerts a strong effect on biological rhythms that is captured in blood by CA, and that this effect is potentially modifiable. This highlights the opportunity to incorporate CA as a measure in precision chronotherapy to evaluate treatment response (optimising dosing time for medications (*49*, *50*), immunotherapy (*51*)), or to quantify the impact of night shift work, unemployment and disability to inform occupational health policy (*52*).

Associations between extreme CA values and mood disorders are consistent with longstanding links between circadian disruption and psychiatric vulnerability (*53*, *54*). However, incident disease associations were modest, and CA was derived from a single time point. Circadian misalignment likely comprises both transitory and chronic components; without repeated measurements, we cannot determine whether elevated CA reflects temporary perturbation or accumulated circadian strain. Again, expanded proteomic datasets with repeated sampling will allow separation of stable trait-like misalignment from transient state effects.

A few limitations should be noted. UK Biobank sampling was restricted to daytime hours, so although inferred nocturnal phase structure was supported by independent 24-hour datasets, overnight trajectories were not directly observed within the cohort itself. CA is also a population-relative measure of circadian alignment rather than an absolute phase marker such as DLMO, and future work should validate it directly against DLMO or against DLMO inferred from wearable accelerometry data in cohorts where this is available. Finally, UK Biobank is an older cohort, so age-related shifts in circadian timing and attenuation of sex differences, including around menopause, may limit generalisability to younger populations.

Our findings establish a scalable framework for molecular circadian phenotyping in population cohorts. Future work should validate proteomic phase prediction against the current field standard of DLMO, examine longitudinal trajectories of CA in disease progression, and test whether CA-guided chronotherapy improves treatment outcomes. We predict that, as biobank-scale proteomics becomes increasingly widespread, molecular circadian phenotyping may become a routine component of epidemiology and precision medicine.

## Acknowledgements

This work was carried out under UK Biobank application 116122. We thank UKB participants and the UKB study team. The chief acknowledgement from CKB is to the participants, the project staff, and the China CDC and its regional offices for assisting with the fieldwork. We thank Judith Mackay in Hong Kong; Yu Wang, Gonghuan Yang, Zhengfu Qiang, Lin Feng, Maigeng Zhou, Wenhua Zhao, and Yan Zhang in China CDC; Lingzhi Kong, Xiucheng Yu, and Kun Li in the Chinese Ministry of Health for assisting with conduct and organization of the study. The CKB baseline survey and the first re-survey were supported by the Kadoorie Charitable Foundation in Hong Kong. The long-term follow-up and subsequent resurveys have been supported by Wellcome grants to Oxford University (212946/Z/18/Z, 202922/Z/16/Z, 104085/Z/14/Z, 088158/Z/09/Z) and grants from the National Natural Science Foundation of China (82192901, 82192904, 82192900) and from the National Key Research and Development Programme of China (2016YFC0900500). The UK Medical Research Council (MC_UU_00017/1, MC_UU_12026/2, MC_U137686851), Cancer Research UK (C16077/A29186, C500/A16896) and British Heart Foundation (CH/1996001/9454), provide core funding to the Clinical Trial Service Unit and Epidemiological Studies Unit, Oxford University for the project. The proteomic assays were supported by BHF (FS/18/23/33512), Novo Nordisk, Olink, SomaScan and NDPH. DNA extraction and genotyping were supported by GlaxoSmithKline and the UK Medical Research Council (MC-PC-13049, MC-PC-14135). We want to acknowledge the participants and investigators of the FinnGen study. The FinnGen project is funded by two grants from Business Finland (HUS 4685/31/2016 and UH 4386/31/2016) and the following industry partners: AbbVie Inc., Alnylam Pharmaceuticals, Inc., AstraZeneca UK Ltd, Bayer AG, Biogen MA Inc., Boehringer Ingelheim International GmbH, Bristol Myers Squibb Inc. (and Celgene Corporation & Celgene International II Sàrl), Genentech Inc., GlaxoSmithKline Intellectual Property Development Ltd., Johnson&Johnson Innovative Medicine Inc., Maze Therapeutics Inc., Merck Sharp & Dohme LCC, Novartis AG, Pfizer Inc. and Sanofi US Services Inc. Following biobanks are acknowledged for delivering biobank samples to FinnGen: Auria Biobank (www.auria.fi/biopankki), THL Biobank (www.thl.fi/biobank), Helsinki Biobank (www.helsinginbiopankki.fi), Biobank Borealis of Northern Finland (https://www.ppshp.fi/Tutkimus-ja-opetus/Biopankki/Pages/Biobank-Borealis-briefly-in-English.aspx), Finnish Clinical Biobank Tampere (www.tays.fi/en-US/Research_and_development/Finnish_Clinical_Biobank_Tampere), Biobank of Eastern Finland (www.ita-suomenbiopankki.fi/en), Central Finland Biobank (www.ksshp.fi/fi-FI/Potilaalle/Biopankki), Finnish Red Cross Blood Service Biobank (www.veripalvelu.fi/verenluovutus/biopankkitoiminta), Terveystalo Biobank (www.terveystalo.com/fi/Yritystietoa/Terveystalo-Biopankki/Biopankki/) and Arctic Biobank (https://www.oulu.fi/en/university/faculties-and-units/faculty-medicine/northern-finland-birth-cohorts-and-arctic-biobank). All Finnish Biobanks are members of BBMRI.fi infrastructure (https://www.bbmri-eric.eu/national-nodes/finland/). Finnish Biobank Cooperative -FINBB (https://finbb.fi/) is the coordinator of BBMRI-ERIC operations in Finland.

## Author contributions

Conceptualization: CA, NRW; Methodology: CA, NRW; Software: CA, SA; Investigation: CA, NRW; Formal analysis: CA, RR, BW, LU, DBR, SA; Data curation: CA, RR, BW, LU; Validation: CA, RR, BW, LU, JJC, YZ, DBR, LL, ZC, IM, HMO, IBH, FG, AK, DWR, NRW; Visualization: CA; Resources: RR, BW, LU, DBR, LL, ZC, IM, HMO, AK, NRW; Supervision: CA, FG, DWR, NRW; Project administration: CA, FG, DWR, NRW; Funding acquisition: CA, JJC, IBH, NRW; Writing – original draft: CA; Writing – review & editing: all authors.

## Data and materials availability

Data may be obtained from a third party and are not publicly available. Researchers can apply to use the UK Biobank resource for health-related research that is in the public interest (https://www.ukbiobank.ac.uk/register-apply/). Individual-level genotypes and register data from FinnGen participants can be accessed by approved researchers via the Fingenious portal (https://site.fingenious.fi/en/) hosted by the Finnish Biobank Cooperative FinBB (https://finbb.fi/en/). FinnGen summary statistics are available through https://www.finngen.fi/en. In CKB, non-genetic data (e.g., baseline, resurveys, biomarkers, and disease follow-up) are released periodically to bona fide researchers. Details of the CKB Data Sharing Policy, data release schedules and data request application procedures are available at www.ckbiobank.org. All queries about data access can be made to. The data underlying the TREASURE study are not publicly available because they are part of an ongoing primary publication and will be reported elsewhere. Code to reproduce our analyses is available at https://github.com/ClaraAlbi/omictime. Biomarker phase and amplitude estimates for UKB data will be deposited in Zenodo upon publication. Time-of-day prediction ML model weights will be deposited in Zenodo upon publication. CA GWAS summary statistics will be deposited in the GWAS Catalog upon publication.

