## Supplementary material for "Population-scale molecular reconstruction of human circadian phase from blood biomarkers"

**The PDF file includes:**

**Materials and Methods**
**Supplementary Text**
**Figs. S1 to S20**
**Tables S1 to S2**
**References**

**Other Supplementary Materials for this manuscript include the following:**

**Data S1 to S9**

### Materials and Methods

### Study Cohorts

#### UK Biobank

UK Biobank (UKB) participants attended the initial assessment visits between 2006–2010 (instance 0). Blood samples were collected at the 7th station of the visit and were processed centrally. Sampling time was obtained from field ID 3166. Up to seven tubes per participant were collected sequentially; the latest timestamp was used as the representative sampling time.

Samples with collection times before 09:00 (n=1,875) or after 20:00 (n=1,904) were excluded to avoid edge effects. The final analytic sample for time-of-day analyses included 493,493 individuals.

Repeat visits (instance 1: "First repeat assessment visit (2012-13)"; instance 2: "Imaging visit (2014+)"; instance 3: "First repeat imaging visit (2019+)") were used for longitudinal validation where biomarker data were available.

#### Blood biomarkers

For the discovery part of our study, we used the available 3,228 blood biomarkers measured on the UKB initial assessment visit (instance 0), including biochemistry, blood cell counts, NMR metabolomics and proteomic (Table S1). More details on quantification procedures and quality control can be found in the following publications: blood biochemistry(Sinnott-Armstrong et al. 2021), blood cell counts(Astle et al. 2016), NMR metabolomics(Julkunen et al. 2023) and proteomics(Sun et al. 2023). Overall, individuals were excluded if they had missing values in time-of-day, self-reported fasting > 24 hours or categorical covariates with <50 observations in any level. Some technical covariates were binned into 20 quantiles and treated as categorical variables (assay date, dilution factor and acquisition time).

Biochemistry: We used 30 available blood biochemistry markers on 469,075 UKB participants. Technical covariates included dilution factor (FID 30897) and date assay (Cat. FID 18518). Raw values were log transformed before analysis.

Blood cell counts: We used 31 available blood haematological assays (counts and percentages) on 476,952 UKB participants. Technical covariates included device ID and time between sampling and processing (Cat. 9081). We excluded individuals where time after venipuncture > 36h. Raw values were log transformed before analysis.

Metabolomics: We used 251 metabolic profiles quantified on 275,000 UKB participants from the Nightingale Health Phase 2. Technical covariates included spectrometer (FID 23650), Processing batch (FID 20282), time between sample preparation and NMR measurement (FID 23659 - FID 23658). Each metabolite was normalized via a log1p transformation and z-scored; extreme values (>4 SD from the mean) were excluded to limit outlier influence.

Proteomics: The proteomics profiling of 53,016 UKB participants was performed using Olink. The data are available as Normalized Protein eXpression (NPX) values, which are relative abundance values in a log2 scale. NPX values were RINT transformed. Technical covariates included Batch (UKB resource 1016), UKB-PPP Consortium selected participant (FID 30903) and OLINK panel processing date (UKB resource 1019). The Olink quantification was performed in 8 separate batches (0-7). We excluded batch 0 (pilot) from analyses (n = 69). Batches 1-6 contain samples from instance 0 and were processed using Olink Explore 3072. Batch 7 is a combination of samples from instances 0, 2 and 3 and were processed using Olink Explore 1536. We ended up with two sets of overlapping proteomics sub-cohorts:

1) Samples from Olink Explore 3072, containing 2,923 proteins from eight Olink panels (Cardiometabolic, Inflammation, Neurology, Oncology, Cardiometabolic II, Inflammation II, Neurology II and Oncology II). We excluded samples with over 2/3rds of protein values missing, resulting in 44,760 individuals for instance 0. Proteins AMY2B, CST1, CTSS, GLIPR1, NPM1, PCOLCE and TACSTD2 were missing in 90% of all samples, so were also excluded from further analyses.

2) Samples from Olink Explore 1536, containing 1,462 proteins from eight Olink panels (Cardiometabolic, Inflammation, Neurology, Oncology). We excluded samples with over 2/3rds of protein values missing, resulting in 52,103 individuals at instance 0. The proteins CTSS, NPM1, PCOLCE, TACSTD2 were missing in 90% of all samples, so were also excluded from further analyses.

**External Validation Cohorts:**

**TREASURE cohort**

12 healthy women and men aged 18 to 30 were recruited to the study (50% female). Blood samples were collected at 3-hour intervals over a 40-hour period, during a Constant Routine protocol to minimise masking effects from light, sleep, food intake, temperature, time awareness and physical activity.

Participants were recruited via flyers and online by the Kramer Lab at the Charité - Universitätsmedizin Berlin. Participants underwent a physical examination and informed consent discussion with a physician from the study team. Inclusion criteria were verified, and participants signed the informed consent form. Participants first underwent a 7-day entrainment phase to ensure participants followed a regular sleep-wake schedule with sufficient sleep before entering the sleep laboratory, where they spent the following 3.5 days. The first night in the sleep lab allowed participants to acclimatize and to detect and exclude participants with previously unknown sleep disorders. On the evening of Day 2, participants were prepared for the upcoming protocol with a peripheral venous catheter inserted. On the morning of Day 3, the 40-hour wakefulness phase began. 14 venous blood draws (via catheter) were conducted according to Charité standard procedures, exclusively by physicians or delegated medical staff. During this period participants remained awake but stayed in bed at all times. Supervision was continuous via video feed to a monitoring room, with staff entering the room if the participant became drowsy. Conditions during the protocol were dim light (<5 lux), stable room temperature and humidity, no access to time cues, hourly iso-caloric snacks (2000 kcal/24h) and water. At the end of the 40-hour wake period, the catheter was removed and the participants were allowed a full night of recovery sleep.

Proteomics: The proteomics profiling of 14 samples from 12 participants (167 samples, NB one sample missing from one participant) was conducted on the Olink Explore HT which assays ~5,400 proteins.

#### China Kadoorie Biobank (CKB)

The China Kadoorie Biobank (CKB) is a prospective cohort study with >512,000 adults aged 30-79 years at baseline(Chen et al. 2011). The participants were recruited during 2004-08 from 10 geographically diverse areas in China. At baseline and subsequent resurveys, participants were assessed by laptop-based questionnaires on socio-demographic characteristics, medical history, and lifestyle habits, with data on physical measurements (e.g. anthropometry, blood pressure, heart rate, lung function) also collected. Non-fasting blood samples were collected, processed, aliquoted, and then stored in liquid nitrogen. After the initial baseline survey, the long-term health of the participants was monitored by linkage with local death or disease registries and with the national health insurance systems.

Proteomics: The proteomics data analysed here were from a subset of 3,977 CKB participants. Protein abundance was measured by the Olink Explore 3072 platform, including a total of 2,941 protein measures. Since six proteins were duplicated across four panels of the Olink platform and showed high correlations (r>0.8), we only kept one measure for each duplicated protein, resulting in 2,923 unique protein abundance measures. Participants with a recorded time earlier than 8 am or later than 9 pm were excluded, resulting in a final sample of 3,965 participants.

#### FinnGen

FinnGen is a public-private research partnership, combining digital healthcare data and multi-omic data on approximately 500,000 Finns (https://www.finngen.fi/en) with the aim of providing novel medical and therapeutic insights into human diseases. FinnGen is a pre-competitive partnership of Finnish biobanks, universities, university hospitals, international pharmaceutical industry partners, and the Finnish biobank cooperative (FINBB). All FinnGen partners are listed at https://www.finngen.fi/en/partners.

Proteomics: The proteomics data presented in this manuscript was collected as part of a pilot study in collaboration with the Finnish Blood Service Biobank, where samples were collected from blood donors who were carriers of specific genetic variants of interest (n=1,990). Data was generated with the Olink Explore 3072 platform(Honkanen et al. 2025). Following QC, individuals missing donation time information (n=714) and whose blood donation was later that 14h (n=11) were removed, resulting in a final sample size of n=1,265. The blood for the majority of individuals in this data was collected between approximately 11h-14h, reflecting the opening hours of Finnish Red Cross Blood Service donation centers and the usual visit time of donors.

### Methods

#### Analysis of variance

To quantify the proportion of variance in biomarker levels attributable to technical, demographic, and genetic factors, we performed analysis of variance (ANOVA) using the aov() function in R (version 4.3.2).

We selected the following covariates: sex (categorical, FID 31), age at recruitment (FID 21002), principal components derived from genome-wide genetic data PC1-20 (FID 22009), assessment centre (categorical, FID 54), self-reported fasting hours (FID 74), month assessment (categorical, FID 55), BMI (FID 21002 / (FID 50/100)^2) and smoking status (categorical, FID 20116, smoking code –3 was set to missing). An interaction term between sex and age at recruitment was also included to capture potential sex-dependent age effects. The variance explained by the sex × age interaction was equally allocated between sex and age categories.

For each biomarker, we modeled the biomarker levels as the outcome variable and included the covariates in the same order:

$$Y_i = \beta_0 + \beta_1 \cdot Sex_i + \beta_2 \cdot Age_i + \beta_3 \cdot BMI_i + \beta_4 \cdot Fasting_i + \beta_5 \cdot AssessmentCentre_i + \beta_6 \cdot Month_i + \Sigma (\beta_k \cdot PC_{ki}) + \beta_t \cdot TechnicalCovariates_i + \varepsilon_i$$

where:

- $Y_i$ = normalized biomarker value for individual i
- $PC_k$ = genetic principal components (k = 1 to 20) provided by the UKB.
- Technical Covariates include batch, processing date, device ID, dilution factor, or other platform-specific variables
- $\varepsilon_i$ = residual error

Partial $R^2$ values were calculated as the ratio of sum of squares for each covariate term to total sum of squares, providing quantitative estimates of relative covariate importance.

**Covariate adjustments**

Biomarker levels were fit using linear regression against different levels of adjustments: no adjustment (no-cov-adjustment), technical adjustment (only quantification platform-specific covariates) and all covariates (excluding time-of-day, BMI and smoking status). The residuals from these models were used in downstream analyses, with each result section explicitly stating which covariate adjustment was used each time.

**Harmonic Regression Modeling**

Circadian rhythmicity was modeled using a first-order harmonic (cosinor-equivalent) regression assuming a 24-hour period(Cornelissen 2014):

$$Y_i = \beta_0 + \beta_1 \cdot \sin(2\pi \cdot t_i / 24) + \beta_2 \cdot \cos(2\pi \cdot t_i / 24) + \varepsilon_i$$

where:

- $t_i$ = sampling time in hours (continuous)
- 24 = assumed circadian period (hours)
- $\beta_1$ and $\beta_2$ = harmonic coefficients

Amplitude (A) was calculated as:

$$A = \sqrt{(\beta_1^2 + \beta_2^2)}$$

Acrophase (φ), expressed in clock hours (0–24), was calculated as:

$$\varphi = (24 / (2\pi)) \times \text{arctangent2}(-\beta_1, \beta_2)$$

Biomarkers were analysed using the all covariate adjustment strategy described above. Models were fitted using the lm function in R (version 4.3.2). Biomarkers were considered rhythmic when the minimum p-value between the sine and cosine harmonic coefficients ($\beta_1$ and $\beta_2$) had an FDR < 5%.

Time x sex interaction models

To test for sex differences in amplitude or phase, nested models were fitted.

Baseline model:

$$Y_i = \beta_0 + \beta_1 \cdot \sin(2\pi \cdot t_i / 24) + \beta_2 \cdot \cos(2\pi \cdot t_i / 24) + \beta_3 \cdot \text{Sex}_i + \beta_4 \cdot \text{Age}_i + \beta_5 \cdot \text{BMI}_i + \beta_6 \cdot \text{Fasting}_i + \beta_7 \cdot \text{AssessmentCentre}_i + \beta_8 \cdot \text{Month}_i + \Sigma\ (\beta_{9+k} \cdot PC_{ki}) + \beta\_T \cdot \text{TechnicalCovariates}_i + \varepsilon_i$$

Interaction model:

$$Y_i = \beta_0 + \beta_1 \cdot \sin(2\pi \cdot t_i / 24) + \beta_2 \cdot \cos(2\pi \cdot t_i / 24) + \beta_3 \cdot \text{Sex}_i + \beta_4 \cdot \text{Sex}_i \cdot \sin(2\pi \cdot t_i / 24) + \beta_5 \cdot \text{Sex}_i \cdot \cos(2\pi \cdot t_i / 24) + \beta_6 \cdot \text{Age}_i + \beta_7 \cdot \text{BMI}_i + \beta_8 \cdot \text{Fasting}_i + \beta_9 \cdot \text{AssessmentCentre}_i + \beta_{10} \cdot \text{Month}_i + \Sigma\ (\beta_{11+k} \cdot PC_{ki}) + \beta\_T \cdot \text{TechnicalCovariates}_i + \varepsilon_i$$

Biomarkers were analysed using the technical covariate adjustment strategy described above. Amplitude and acrophase were calculated as described above. Models were fitted using the lm function in R (version 4.3.2). Biomarkers were considered to have sex x time specific effects when the p-value from an ANOVA F-test between the nested models had FDR < 5%.

Longitudinal Harmonic Modeling

For repeated-measure datasets (TREASURE), mixed-effects cosinor models were fitted:

$$Y_{ij} = \beta_0 + \beta_1 \cdot \sin(2\pi \cdot t_{ij} / 24) + \beta_2 \cdot \cos(2\pi \cdot t_{ij} / 24) + b_{0j} + \varepsilon_{ij}$$

where:

- j indexes participants
- $b_{0j}$ = participant-specific random intercept
- $\varepsilon_{ij}$ = residual error

Amplitude and acrophase were calculated as described above. Models were fitted using the lme4 function in R (version 4.3.2).

Acrophase and amplitude correlations with external datasets

Correlations between acrophase sets were estimated using Pearson's correlation coefficient from cor.test function in R (version 4.3.2). To account for phase circularity, all phase estimates that had a UKB equivalent with >12h difference were projected onto the shortest 24-hour distance (−12 to +12 h) to account for wrapping across midnight. This resulted in acrophase values >24h.

We calculated correlations using top 86 rhythmic proteins out of 131 rhythmic biomarkers ($R^2$>1%, amplitude>0.1 in UKB). The TREASURE cohort is described in the section above, where proteins PLAT and GIP were missing from the Explore HD panel. The SomaScan harmonic estimates were taken from Specht *et al.(Specht et al. 2024)*, where plasma samples from 17 participants (33% female) were collected under controlled and regular sleep-wake schedule conditions. We matched the proteins by UniProt, obtaining 53 SomaScan analogs (44 unique gene names). We restricted comparisons to the harmonic parameters (acrophase and amplitude) from the 1st order harmonic models (variables "acrophase of 1-harmonic fit (time to DLMO in hours)" and "amplitude of 1 harmonic fit" from the supplementary Table 1).

**Proteomic tissue of origin analyses**

In order to classify the Olink plasma proteins with a specific tissue of origin, we used data from the "Tissue resource - Tissue-based map of the human proteome" from The Human Protein Atlas (https://www.proteinatlas.org/). This dataset provides *tissue specificity* measures for each protein based on the expression of the gene in the GTEx resource(GTEx Consortium 2020). Elevated expression includes these subcategory types of elevated expression:

- **Tissue enriched (single):** At least four-fold higher mRNA level in a particular tissue/cell type compared to any other tissues/cell types.
- **Group enriched (group):** At least four-fold higher average mRNA level in a group of 2-5 tissues or 2-10 cell types compared to any other tissues/cell types.

Out of the 1,406 rhythmic proteins, 206 were classified as "Tissue enriched", 303 were "Group enriched", 801 were "Tissue enhanced" and the rest had "Low tissue specificity". We focused on the first two subcategories for robustness.

**Time-stamp algorithms**

A 5-fold cross-validation scheme was used, randomly assigning participants to one of five subsets. Models were trained on 80% of the data and tested on the remaining 20%, iterating through each fold. All biomarker values were standardised prior to modelling.

Four predictive approaches were evaluated: LASSO regression, quadratic LASSO (including squared biomarker terms), and two gradient boosting methods (XGBoost and LightGBM). LASSO was selected to enable feature selection and identify a minimal subset of informative biomarkers, while tree-based models were included to capture potential non-linear relationships and interactions.

Model Specifications

1. **LightGBM.** A gradient-boosting model was trained using the LightGBM R package(Ke et al. 2017). Standard hyperparameters setting included feature and bagging fractions of 0.8, lambda regularizations (L1 = 5, L2 = 2), minimum data per leaf (50), and early stopping with a maximum of 1000 iterations. Optimal iterations were determined using internal 5-fold cross-validation within the training subset.
2. **XGBoost** An XGBoost regression model was trained using the XGBoost R package(Chen and Guestrin 2016) with learning rate (eta) of 0.1, maximum tree depth of 6, and 0.8 subsampling for both data and features. The model was trained for 100 boosting rounds.
3. **LASSO Regression.** A penalized linear regression model using the Least Absolute Shrinkage and Selection Operator (LASSO) was trained using the glmnet R package(Friedman et al. 2010). Missing data were imputed automatically. The optimal lambda parameter was selected via internal cross-validation.
4. **LASSO Regression with Quadratic Terms.** An additional LASSO model was trained, including both linear and quadratic terms (squared biomarkers), to capture potential nonlinear relationships. Quadratic terms were generated by squaring each numerical standardised biomarker.

Missing data were handled according to model requirements: for LASSO models, missing values were mean-imputed; gradient boosting methods natively accommodate missing values during training. For each model, prediction performance was assessed using the coefficient of determination ($R^2$), calculated as the squared correlation between predicted and recorded sampling time in held-out folds.

Platform-specific models (proteomics, metabolomics, biochemistry, cell types) were trained independently. An additional model integrating all biomarkers was evaluated in the subset of participants with complete data across platforms (multi-platform). Predictions from the four algorithms were averaged to obtain final time-of-day estimates for downstream analyses.

All modeling was performed in R (version 4.3.2) using glmnet, xgboost, and lightgbm packages.

UKB repeated blood biomarker measures

We further evaluated the prediction accuracy of the metabolomics and proteomics models in a subset of UKB participants with repeated measures. For metabolomics, 16,665 samples had available Nightingale

Health NMR measurements for the same 251 metabolites also at instance 1 (First repeat assessment visit, 2012-13).

For proteomics, the repeated measures were performed using the Olink Explore 1536 platform, consisting of half the panels of the Olink Explore 3072 platform. After the same QC steps as in the full data, we analysed 1,157 samples at instance 2 (Imaging visit, 2014+) and 1,109 at instance 3 (First repeat imaging visit, 2019+) with 1,457 quantified proteins. We retrained the time prediction models on the instance 0 proteomic dataset, restricting the data to the proteins in Olink Explore 1536 platform. After the same QC steps as in the full data, we obtained 51,719 samples and 1,457 proteins. Again, we used 5-fold cross-validation to train four models: LightGBM, XGBoost, LASSO and LASSO_x2. We used model weights from cross-validation training set 1 and projected them into the NPX protein values of the repeated measures. Used the mean prediction across the four prediction models.

The time-of-day of blood sampling for the repeated samples was extracted for instances 1, 2 and 3 (Field ID 3166) (Fig. S9).

<u>Prediction External Validation</u>

A detailed description of external validation cohorts is available above (CKB, FinnGen and TREASURE). We used model weights from cross-validation training set 1 and projected them into the NPX protein values of the cohorts. Used the mean prediction across the four prediction models.

Prediction accuracy was again assessed using the coefficient of determination ($R^2$), calculated as the squared correlation between predicted and recorded sampling times. In the case of TREASURE, we derived two $R^2$ estimates, one that limited the samples to the 9-20h range to match the UKB ($R^2 = 0.59$) and one that modelled time in a cosinor function assuming a 24h period ($R^2 = 0.66$); i.e., the time of day variation (denominator of R2) changed, as well as the numerator.

**Definition of Circadian Acceleration (CA)**

For downstream analyses, we used the proteomic circadian phase predictor trained on 1,459 proteins (cross-validated $R^2 = 0.68$; n = 51,701). Final predicted time-of-day values were obtained by averaging predictions across the four machine-learning models.

Circadian acceleration (CA) was defined as the residual from a linear regression of predicted time-of-day on recorded clock time:

$\text{PredictedTime}_i = \beta_0 + \beta_1 \cdot \text{RecordedTime}_i + \varepsilon_i$

$\text{CA}_i = \varepsilon_i$

where:

- $\text{PredictedTime}_i$ is the model-inferred circadian phase (in hours) for individual i

- $RecordedTime_i$ is the observed clock time of blood sampling
- $\varepsilon_i$ represents the deviation from the population-average phase at that clock time

Thus, CA captures the extent to which an individual's inferred circadian phase is advanced (positive residual) or delayed (negative residual) relative to the expected phase for that sampling time.

By construction, CA is centered at zero across the population and represents a population-relative measure of circadian phase misalignment rather than absolute biological phase.

Longitudinal Stability of Circadian Acceleration

For UKB participants with repeated proteomic measurements (up to 3), CA was calculated independently at each visit. Pairwise correlations of CA across visits were computed to assess temporal stability. For comparison, correlations between recorded sampling times across visits were also calculated.

To assess within- and between-individual variability in circadian acceleration (CA), we fitted linear mixed-effects models including a participant-specific random intercept:

$$CA_{ij} = \beta_0 + \beta_1 \cdot RecordedTime_{ij} + b_{0j} + \varepsilon_{ij}$$

where $b_{0j}$ represents stable between-individual differences and $\varepsilon_{ij}$ residual within-individual variability. Models were implemented using the lmer() function (lme4 package in R).

Long-term stability was quantified using the intraclass correlation coefficient (ICC), calculated as the proportion of variance attributable to the participant-level random effect:

$$ICC = \sigma^2\_between / (\sigma^2\_between + \sigma^2\_within)$$

ICC values were computed using the performance package in R (version 4.3.2).

### Phenotypic associations

All analyses were performed in 51,719 UKB participants in whom the proteomics-based circadian acceleration (CA) measure was derived. All phenotypic outcomes were measured at the initial assessment visit, concurrent with blood sample collection. Sample sizes for individual variables are available in Data S3.

Sociodemographic covariates included age at recruitment (FID 21002), analysed as a continuous variable, sex (FID 31), body mass index calculated from measured weight (FID 21001) and standing height (FID 12144), smoking status (FID 20116; never, former, current), and socioeconomic deprivation assessed using the Townsend deprivation index (FID 189). Genetic ancestry (FID 30079) was included as a categorical variable, with European ancestry as the reference group(Karczewski et al. 2024).

Sleep-related variables were obtained from the baseline self-report questionnaires and included chronotype (FID 1180), average sleep duration (FID 1160, categorised into short (<7 h), normal (7–9 h), or long (>9 h) in accordance with established guidelines(Kyle et al. 2017)), insomnia symptoms (FID 1200), and ease of waking (FID 1170).

The UKB online followup sleep questionnaire (2023) was used to compare the single-item chronotype question (FID 30429, at baseline FID 1180) and the Reduced Morningness–Eveningness Questionnaire (rMEQ)(Adan and Almirall 1991) a validated 5-item self-report measure of diurnal preference (FIDs 30425 - 30429), used to classify participants along the morningness–eveningness continuum, which was then categorised into 5 chronotypes for comparison, based on the guidelines.

Employment-related variables included current employment (FID 6142), shift work frequency (FID 826) and night shift work frequency (FID 3426), as well as an interaction variable combining chronotype (morning vs evening preference) with night shift work status.

Season-related variables were derived from the blood collection timestamp (FID 3166). Season of sampling (season) was derived from the month of blood collection and categorised as Winter (December–February), Spring (March–May), Summer (June–August), or Fall (September–November). Social jetlag was evaluated with the day-type variable (day_type) classifying samples as weekday or weekend, with weekends defined as samples collected on Saturdays. A categorical indicator (fri_sun) was created to identify samples collected on Friday, Saturday, or Monday. Daylight saving time status (is_dst) at sampling was determined using the Europe/London timezone. Proximity to daylight saving time transitions was additionally captured using separate categorical variables for spring (springDST) and autumn (autumnDST), classifying samples as baseline (3–14 days before), immediately before (1–2 days before), or immediately after (1–2 days after) the transition.

Self-reported medication use was mapped to ATC codes from FID 20003 using the table in (Wu et al. 2019). We selected medications that affect sleep as described in(Kyle et al. 2017), namely antidepressants (N06A), mood stabilizers (N03AG01, N03AX09, N03AF01, N03AF02, N05AN01), hypnotics/sedatives (N05C), and antihypertensive medication including ACE inhibitors (C09A), angiotensin II receptor blockers (C09C), beta blockers (C07), calcium channel blockers (C08), and diuretics (C03).

**Disease outcomes**

Disease outcomes were ascertained using UKB derived fields capturing the date of first occurrence of diagnoses from linked hospital inpatient, primary care, and death registry records (UKB Category 1712). Participants were classified as affected if a diagnosis was recorded, with the earliest available date used as the onset. Autoimmune disease included rheumatoid arthritis (M05/M06; FIDs 131848/131850). Cardiometabolic conditions included hypertension (I10; FID 131286), dyslipidaemia (E78; FID 130814), chronic ischaemic heart disease (I25; FID 131306), obesity (E66; FID 130792), and type 2 diabetes mellitus (E11; FID 130708). Neuropsychiatric disorders included depression (F32/F33; FIDs 130894/130896), bipolar affective disorder (F31; FID 130892), schizophrenia (F20; FID 130874), and dementia (F03; FID 130842). We did not include sleep disorders (G47) because of the low number of cases in the UKB proteomics subset.

Prevalent disease outcomes were defined using a 15-year lookback window prior to baseline. For each condition, participants were classified as prevalent cases if the first recorded diagnosis occurred within the 15 years preceding the blood sampling date (including the baseline date). Participants with no recorded diagnosis were classified as non-cases, while diagnoses occurring more than 15 years before baseline were excluded from the analysis.

Incident disease outcomes were defined using a 15-year follow-up window after baseline. Participants were classified as incident cases if the first recorded diagnosis occurred after the blood sampling date and within 15 years of follow-up. Participants with no recorded diagnosis during this period were classified as non-cases, while diagnoses occurring prior to baseline or beyond 15 years after baseline were excluded.

Participants were additionally classified into three groups based on their CA levels: Accelerated (> +2 CA), Delayed (< −2 CA), and Middle (−2 to +2 CA), with the Middle group serving as the reference. Baseline characteristics were summarised by group (Data S4). Group differences were assessed using pairwise comparisons of Accelerated versus Middle and Delayed versus Middle. Continuous variables were compared using two-sample t-tests or Wilcoxon rank-sum tests as appropriate, and categorical variables using chi-squared or Fisher's exact tests. All tests were two-sided and p-values were reported.

### Statistical analyses

Associations between circadian acceleration (CA) and continuous phenotype outcomes were tested using linear regression in R, implemented using the lm() R function (version 4.3.2), with CA included as the main exposure variable. All phenotype models were adjusted for sex, age at recruitment, UKB assessment centre, and the first 20 genetic principal components.

Associations between circadian acceleration (CA) and prevalent disease outcomes were tested using logistic regression in R, with CA included as the exposure of interest. Associations with incident disease outcomes were tested analogously using logistic regression restricted to the incident outcome definitions. Models were adjusted for sex, age at recruitment, chronotype, season of blood sampling, socioeconomic deprivation, BMI, smoking status, UKB assessment centre, and the first 20 genetic principal components.

### Genetic analyses

UKB data

Genetic analyses were conducted in UKB participants of inferred European ancestry. Ancestry was defined based on categories provided by the UKB in FID 30079(Karczewski et al. 2024).

For GWAS analyses, we included 44,351 unrelated individuals of European ancestry with available circadian acceleration (CA) estimates. For SNP-based heritability analyses, we restricted to a subset of 30,327 unrelated individuals with a relatedness cutoff of 0.05. We constructed a sparse genomic relationship matrix (GRM) for the full set of UKB participants using GCTA (v1.95)(Yang et al. 2011, 2010) and restricting to the HapMap set of variants.

UKB imputed genotype data (v2), resulting in 8,545,378 bi-allelic genetic variants with minor allele frequency (MAF $\geq$ 0.01).

Genome-wide association study (GWAS)

Genome-wide association analysis was performed using fastGWA(Jiang et al. 2019), implemented in GCTA (v1.95). fastGWA uses a sparse GRM to account for relatedness and population structure.

We performed a GWAS of CA and a GWAS of chronotype, where the self-reported answers were transformed into a quantitative trait (−2 = definite evening to +2 = definite morning) following Jones *et al.* 2019 (Jones, Lane, et al. 2019). Covariates included sex, age, 20 first genetic principal components, genotype batch and UKB assessment centre.

Chromosome X association analyses were conducted separately in males and females and meta-analysed using fixed-effect inverse-variance weighted (IVW) meta-analysis.

Independent association signals were identified using GCTA-COJO on each chromosome separately (--cojo-slct). Variants selected by COJO were considered conditionally independent signals. Functional annotation of lead variants was performed using ANNOVAR("ANNOVAR: Functional Annotation of Genetic Variants from next-Generation Sequencing Data Nucleic" 2010).

Fine-mapping

We identified putative causal variants for CA using PolyFun(Weissbrod et al. 2020) and FINEMAP v1.4.1(Benner et al. 2016). We restricted the summary statistics to variants lying outside of the HLA region. We computed prior causal probabilities via the L2-regularized extension of S-LDSC described in Weissbrod et al. using the baseline LF2 genome annotations. We then performed fine-mapping on the genome-wide significant loci (pvalue < 5e-8) by 3Mb windows. We used FINEMAP to perform fine-mapping, assuming at max. 5 causal variants. We identified variants of interest as having a posterior inclusion probability in the causal set (PIP) > 0.95. We ranked all variants by PIP and defined 95% credible causal sets of variants as the minimum set of variants whose PIPs summed to >=0.95.

Stratified LD score regression (S-LDSC) (Finucane et al. 2015) was used on the GWAS summary statistics to identify tissue-specific heritability enrichments.

Genetic correlations

Genetic correlations were estimated using LDSC(Bulik-Sullivan et al. 2015). We estimated pairwise genetic correlation between CA, chronotype and 9 accelerometer-derived sleep and activity measures(Jones, van Hees, et al. 2019; Ferguson et al. 2018):

- Mean diurnal inactivity duration (ACC_DIURNAL_INACT): Average duration of inactivity occurring outside the sleep period time (SPT) window.
- Mean L5 timing (ACC_L5_TIME): Mean timing of the least active 5-hour period (L5), expressed as hours elapsed since the previous midnight.

- Mean M10 timing (ACC_M10_TIME): Mean timing of the most active 10-hour period (M10), expressed as hours elapsed since the previous midnight.
- Mean number of sleep episodes (ACC_N_SLEEP_EPISODES): Average number of discrete sleep episodes occurring within the SPT window, adjusted for mean SPT-window duration.
- Mean sleep duration (ACC_SLEEP_DUR): Average duration of sleep occurring within the SPT window.
- Sleep duration variability (ACC_SLEEP_DUR_SD): Standard deviation of sleep duration across all recorded SPT windows.
- Mean sleep efficiency (ACC_SLEEP_EFF): Average proportion of the SPT window classified as sleep.
- Mean sleep midpoint (ACC_SLEEP_MIDP): Average midpoint of the SPT window, expressed as hours elapsed since the previous midnight.
- Relative amplitude (RA). *RA*= (ACC_M10_TIME−ACC_L5_TIME) / (ACC_M10_TIME+ACC_L5_TIME)

We also estimated genetic correlations between CA and 16 selected circadian-related traits and diseases: **Insomnia** (P. R. Jansen et al. 2019), **MDD** (3 phenotypes)(Major Depressive Disorder Working Group of the Psychiatric Genomics Consortium. Electronic address: and Major Depressive Disorder Working Group of the Psychiatric Genomics Consortium 2025), **Schizophrenia**(Trubetskoy et al. 2022), **Bipolar disorder** (O'Connell et al. 2025), **Coronary artery disease** (Aragam et al. 2022), **T2D** (Suzuki et al. 2024), **Rheumatoid arthritis**(Ishigaki et al. 2022), **mtDNA**(Longchamps et al. 2022), **BMI**(Locke et al. 2015), **BMI-adjusted hip circumference**(Christakoudi et al. 2021), **Fasting glucose**(Lagou et al. 2023), **Hypertension, Diastolic Blood Pressure**(Surendran et al. 2020) and **Alzheimer's disease**(I. E. Jansen et al. 2019).

<u>Mendelian Randomisation</u>

Bi-directional two-sample Mendelian randomisation (MR) was performed in R using the TwoSampleMR package (v0.6.27)(Hemani et al. 2018). For each exposure–outcome pair, causal effects were estimated using inverse-variance weighted MR under a multiplicative random-effects model, alongside weighted median and MR-Egger regression as sensitivity analyses. Between-instrument heterogeneity was assessed using Cochran's Q. Directional horizontal pleiotropy was evaluated using the MR-Egger intercept. Outlier instruments and outlier-driven horizontal pleiotropy were assessed using MR-PRESSO (global and outlier tests, with distortion testing where applicable).

<u>Polygenic Scores</u>

We derived a CA polygenic score (PGS) using SBayesRC(Zheng et al. 2022). We re-estimated the CA GWAS excluding 1,052 individuals of European ancestry with repeated measures from the GWAS sample to obtain an out-of-sample prediction accuracy of the PGS.

The PGS weights were projected into the left-out UKB data without proteomics, resulting in 381,449 unrelated individuals. We validated the CA PRS by estimating the relationship between the CA PRS and self-reported chronotype; including sex, age and 20 PCs as covariates.

### Supplementary Text

#### ST1. Functional classification of rhythmic blood biomarkers

We highlight strong time-of-day variation in routine clinical biomarkers across multiple blood cell counts (leukocytes, lymphocytes, monocytes, neutrophils, basophils), as well as standard biochemistry biomarkers (phosphate and total bilirubin).

The proteomic results included inflammatory and immune mediators (IL6, CXCL8, CCL3, TNFSF14, CD40LG, GZMB, SLAMF8, RNASE6, TCL1A, TREML1), growth factors and endocrine regulators (EGF, FGF19, GDF15, GIP, GCG, PRL, PTH, INHBB, FST, ADM, AGRP), angiogenic and vascular factors (ANGPTL1, ANGPTL2, ANGPTL4, EDN1, PGF, SEMA3F, SEMA4D), extracellular matrix and remodeling proteins (ADAMTS15, MFAP4, MFGE8, SMOC1, SPON2, TIMP4, HYAL1, SERPINE1, SDC1, SBSN), proteases and processing enzymes (CPA2, CPA4, CTSE, FURIN, KLK13, MEP1A, PCSK9, PLAT, TMPRSS15, GLB1, HS3ST3B1), adhesion and receptor molecules (CD84, CDH1, EPHA1, TNFRSF11B, TNFRSF12A, SUSD1, TNR), and structural or intracellular regulators (ACTN2, MYBPC1, MYL3, HSPB6, LSP1, TMSB10, SF3B4, VTA1).

Small-molecule metabolites included amino acids (alanine, phenylalanine, leucine, isoleucine, valine, and total branched-chain amino acids), glycolytic intermediates (glucose, lactate), ketone bodies (3-hydroxybutyrate, acetoacetate, acetone), acetate, triglycerides, and monounsaturated fatty acids. The lipoprotein profiling captured HDL, LDL, and VLDL particle size, concentration, and compositional measures (triglycerides, phospholipids, free cholesterol, and cholesteryl esters), reflecting lipid transport and remodeling.

#### ST2. Platform-Specific and Combined Model Performance

Prediction performance varied across biomarker platforms and modeling approaches (Fig. S6). For each platform, final predictions were obtained by averaging outputs from four algorithms (LASSO, quadratic LASSO, XGBoost, and LightGBM). Proteomics achieved the highest mean cross-validated accuracy ($R^2$ = 0.68), followed by metabolomics ($R^2$ = 0.42), clinical biochemistry ($R^2$ = 0.17), and hematological traits ($R^2$ = 0.15).

Within proteomics, the highest-performing individual model was quadratic LASSO ($R^2$ = 0.68). For metabolomics, biochemistry, and hematology, LightGBM yielded the strongest performance. Predictions generated by different algorithms within the same platform were highly correlated (Pearson's r = 0.86–0.98; Fig. S7), indicating that performance reflected shared temporal signal rather than algorithm-specific effects. For downstream analyses, we used the averaged proteomic predictor.

When all 3,228 biomarkers were modeled jointly in the subset of individuals with complete multi-platform data (n = 22,461), prediction accuracy increased modestly to $R^2$ = 0.71 (Fig. S3A). Cross-platform predictions were moderately correlated (Pearson's r = 0.26–0.63; Fig. S8), while the combined model showed strong correlation with proteomics (r = 0.96) and metabolomics (r = 0.74), indicating substantial overlap in circadian information captured by these molecular layers. Given the modest performance gain relative to proteomics alone and the reduction in sample size, subsequent analyses were conducted using the proteomic model.

Proteomic measurements were generated across multiple Olink batches. Batches 1–6 (Explore 3072) included instance 0 samples with 2,916 proteins quantified, while Batch 7 (Explore 1536) included a mixture of instance 0, 2, and 3 samples with 1,462 proteins measured. To maximize comparability across visits and batches, models were retrained using the 1,458 proteins shared across platforms. Restricting to this common protein set yielded equivalent predictive performance ($R^2$ = 0.68) while increasing the analyzable baseline sample size to 51,701 individuals by incorporating Batch 7 samples. These results indicate that predictive performance plateaued below the full 2,916-protein panel and was not dependent on the larger protein set.

**ST3. Covariate Adjustment Sensitivity**

To evaluate the robustness to the covariate adjustment strategy, models were trained under three adjustment schemes: (1) no adjustment, (2) adjustment for platform-specific technical covariates only, and (3) full biological and technical covariate adjustment (excluding time-of-day, BMI, and smoking status). This comparison was conducted in the subset of 51,701 participants with 1,459 shared proteins. Prediction performance was stable across adjustment strategies, with mean $R^2$ values of 0.69 (no adjustment), 0.68 (technical adjustment), and 0.68 (full adjustment) (Fig. S12), indicating that time-of-day prediction was not materially influenced by covariate preprocessing.

We assessed potential sex differences in prediction accuracy by training models separately in males and females using the technical-adjusted data. Mean prediction accuracy was slightly higher in females ($R^2$ = 0.68) than in males ($R^2$ = 0.65), consistent with minor differences in sample size and/or rhythmic amplitude between sexes.

**S4. Feature Reduction and Translational Feasibility**

To evaluate translational feasibility of blood-based circadian time inference—including potential clinical and forensic applications—we assessed whether prediction accuracy could be retained using reduced biomarker panels. Using the LASSO model trained on 1,459 proteins in 51,701 participants, model coefficients were thresholded at increasing values (t = 0, 0.01, 0.05, 0.1), retaining progressively smaller subsets of proteins.

Prediction performance decreased gradually with increasing threshold stringency. Mean cross-validated $R^2$ was 0.66 at t = 0 (1,132 proteins retained), 0.66 at t = 0.01 (759 proteins), 0.58 at t = 0.05 (215 proteins), and 0.40 at t = 0.1 (66 proteins) (Fig. S13). Feature counts were averaged across the five cross-validation folds. These results indicate that substantial prediction accuracy can be maintained with markedly reduced protein panels, supporting feasibility of compact assays for applications where measuring the full high-dimensional proteome is impractical. The UKB has announced an upcoming expansion of Olink proteomics to the full ~500,000 cohort. When available, this increase in training sample size is expected to improve estimation of lower-amplitude rhythmic signals and enable more stable model fitting under feature reduction, thereby increasing power to develop smaller targeted protein panels without substantial loss of predictive performance.

### S5. Extended Phenotypic Associations and Disease Analyses of Circadian Acceleration

S5.1 Phenotypic characterisation of circadian acceleration (CA)

We tested associations between circadian acceleration (CA) and socio-demographic, environmental, and self-reported variables collected at the UKB assessment (Data S3). CA showed no association with age, sex, genetic ancestry, smoking status, or Townsend Deprivation Index (TDI). BMI showed a statistically significant but negligible effect size (corresponding to <1 min shift). We note that blood proteins used for phase prediction were pre-adjusted for sex, age, genetic principal components, and technical covariates, which may attenuate associations of these variables with CA.

Seasonal and social timing effects were assessed using blood collection date and appointment day (Fig. 4C). Relative to winter, CA was delayed in summer (+6.6 min) and spring (+3.3 min). To quantify weekend-related effects, we compared Saturday appointments to weekday appointments (no Sunday appointments were available). Saturday appointments were delayed by 3.6 min relative to weekdays. For daylight saving time (DST), we compared CA in the days −2 to −1 before and +1 to +2 after the spring and autumn DST weekends against a baseline defined as days −14 to −3 before DST. No significant change was observed for the autumn transition; in contrast, CA was delayed by 18.8 min on the Monday-Tuesday after spring DST (Fig. 4C).

Sleep-related phenotypes showed the largest effect sizes (Fig. 6C). Chronotype exhibited a graded pattern, with "Definitely evening" delayed by 37.6 min relative to "Definitely morning," and intermediate categories showing progressively smaller delays ("Rather evening": 24.1 min; "Don't know": 15.6 min; "Rather morning": 9.0 min). Sleep inertia was also associated with delayed CA: participants reporting "Not at all easy" to get up were delayed by 26.3 min relative to "Very easy," with intermediate groups showing graded effects ("Not very easy": 19.6 min; "Fairly easy": 9.6 min). Short sleep duration (<7 h) was associated with a modest shift relative to 7–9 h (3.6 min), while long sleep duration (>9 h) was not significantly different from 7–9 h. No significant association was detected between CA and self-reported insomnia.

We used data from the UKB follow-up online sleep questionnaire (2023) to compare the effect on CA of the single-item chronotype definition and the rMEQ-based chronotype, which is defined by weighting 5 sleep-related questions. The distributions of the two chronotype definitions show how extreme chronotypes ("Definitely morning/evening") were more frequent in the single-item question, and these get re-defined by the other complementary questions, into the centre "Neither" (Fig. S14A). The single-item chronotype effect on CA remained similar after ~17 years, with "Definitely evening" delayed by 30 min relative to "Definitely morning", highlighting the stability of self-reported chronotype over time. Interestingly, the rMEQ chronotype from the same individuals had much larger effects on CA, with the "Definitely evening" being delayed by 81 min with respect to "Definitely morning" (Fig. S14B). Again, all intermediate chronotypes showed dose responses with proximity to the reference "Definitely morning". These results provide new biological evidence for the validation of rMEQ as a more precise measure of chronotype.

Employment status and shift work were also associated with CA (Fig. 4C). Relative to employed participants, retired participants were delayed by 3.4 min and those reporting disability by 6.2 min. Among employed participants, "always" shift work was associated with a 14.1 min delay relative to "never" shift work. Night shift exposure showed a dose-response pattern: participants consistently working night shifts were delayed by 22.3 min, whereas those who usually (but not always) worked nights were delayed by 14.0 min.

Finally, we assessed whether self-reported use of sleep-related medications (including sedatives and hypnotics; see Methods) was associated with CA. No significant associations were observed.

S5.2 Circadian acceleration as a disease risk factor

We evaluated associations between CA and 12 diseases with established circadian involvement across neuropsychiatric (depression, bipolar disorder, schizophrenia, dementia), cardiometabolic (type 2 diabetes, obesity, hypertension, dyslipidemia, chronic liver disease, chronic heart disease), and immune (rheumatoid arthritis) categories (references as in main text). CA was analyzed as (i) a continuous variable and (ii) categorical extremes: accelerated ($CA > 2$), delayed ($CA < -2$), and middle ($-2 \leq CA \leq 2$).

Initial comparisons of participant characteristics across CA categories showed that the accelerated and delayed tails differed from the middle group by sex, age, ancestry, TDI, and smoking status (Data S4). Tails were enriched for younger males who were current smokers, had lower TDI scores, and were employed or reported disability at assessment. The delayed group also included higher proportions of inferred African and South Asian genetic ancestries. These covariates, together with phenotypic variables previously associated with CA, were included in downstream disease association models.

Prevalent disease (≤15 years before sampling)

To assess associations of CA with existing disease at baseline, we defined prevalent cases as diagnoses occurring within 15 years prior to blood sampling (excluding earlier diagnoses). This window was chosen to align with observed intra-individual CA correlations over similar time scales.

Continuous CA analysis showed decreased odds for dyslipidemia (OR = 0.95; pFDR = $1.88 \times 10^{-3}$) and chronic heart disease (OR = 0.94; pFDR = 0.02) (Fig. S16). No other conditions were significant for continuous CA. In categorical analyses, the accelerated group showed increased odds of episodic depression (OR_accelerated = 1.34; pFDR = $8.89 \times 10^{-3}$) and bipolar disorder (OR_accelerated = 3.04; pFDR = 0.02). Case numbers in extreme groups were limited, particularly for bipolar disorder (n = 11 accelerated).

Incident disease (≤15 years after sampling)

For incident disease analyses, we identified cases diagnosed up to 15 years after blood sampling among individuals without prevalent disease. No associations survived FDR correction for either continuous CA or CA extremes (Fig. S16), although number of incident cases in extreme groups was null for disorders with onset during adolescence and early adulthood (bipolar disorder, schizophrenia). Nominal associations were observed for chronic liver disease (OR_accelerated = 1.27; p = 0.04).

### S6. Genetic Analyses of Circadian Acceleration

#### S6.1 SNP-based heritability

SNP-based heritability of circadian acceleration (CA) was estimated using GCTA-GREML (Yang et al. 2011) in n = 30,327 unrelated participants of inferred European ancestry. The heritability estimate was $h^2 = 0.10$ (SE = 0.01).

#### S6.2 Genome-wide association analysis

Genome-wide association analysis (GWAS) was performed using fastGWA (Jiang et al. 2019) in n = 44,529 participants of inferred European ancestry. A total of 1,825 variants reached genome-wide significance ($P < 5 \times 10^{-8}$) (Fig. 5A).

Conditional and joint analysis using COJO (Yang et al. 2012) identified 20 independent association signals across 11 chromosomes , Including exonic variants in *MYOC* (chr1) and *NLRP12* (chr19), intronic variants in *EFNA1*, *TNR* (chr1), *SYN2* and *DOCK3* (chr3), *SPON2* (chr4), *SPINK5* (chr5) and *COLEC10* (chr8), *FAS* (chr10), *RELT* (chr11), and *GDF15* (chr19); and intergenic and upstream/downstream variants to *ANGPTL1/RALGPS2* and C1orf220 (chr1), *PGF* (chr14), *HS3ST3A1* (chr17), *KLK11/KLK12* (chr19), and *LGALS1* (chr22) (Data S5).

#### S6.3 Fine-mapping and functional enrichment

Fine-mapping was conducted using PolyFun/FINEMAP(Weissbrod et al. 2020; Benner et al. 2016). Five credible sets were identified, including three single-variant credible sets with posterior inclusion probability (PIP) > 0.95: rs34036521 (ANGPTL1, chr1), rs1055150 (GDF15, chr19), and rs34436714 (NLRP12, chr19). Two additional loci (chr17 near HS3ST3A1 and chr19 near KLK11/KLK12) comprised 2–3 variants with cumulative posterior probability > 95% (Data S6).

Stratified LD score regression (S-LDSC) (Finucane et al. 2015) was performed using baseline annotations. No categories showed significant enrichment after multiple-testing correction.

#### S6.4 Overlap with proteins used in phase prediction and pQTL follow-up

Of the 20 COJO lead variants, 15 mapped to genes encoding 13 plasma proteins quantified on Olink Explore and included in the phase prediction models (including two independent signals at ANGPTL1 and GDF15). For HS3ST3A1, a closely related protein (HS3ST3B1) was quantified. This overlap suggests that part of the CA genetic signal is mediated through genetic effects on circulating protein abundance.

We evaluated additive genotype–protein associations for these variants in UKB and observed expected allele-dosage effects on corresponding protein levels (Fig. S17). For the remaining five COJO variants (mapping to DOCK3, SYN2, COLEC10, C1orf220, and NLRP12), we tested whether they were associated with levels of proteins implicated above. Significant associations were observed between rs2761462 (COLEC10 locus) and ANGPTL1 protein levels, rs34436714 (NLRP12 locus) and LGALS1 protein levels, and rs184262 (SYN2 locus) and SPINK5 protein levels (Fig. S18). The remaining variants showed no detectable associations with these proteins in cis; additional trans effects on proteins included in the prediction models cannot be excluded.

Collectively, these results indicate that a subset of CA-associated variants map to proteins that also exhibit strong time-of-day variation, consistent with genetic effects on circulating protein abundance contributing to the CA signal. For example, several loci showed clear additive allele-dosage effects on protein levels (Figs. S15–S16). In most cases, individual genotypes were associated with differences in rhythmic amplitude estimates, with mean amplitude differing by up to ~1 unit between individuals carrying 0 versus 2 copies of the allele (e.g., RELT; Fig. S17).

S6.5 Genetic correlations with chronotype and accelerometer-derived sleep timing

To assess whether CA shares genetic architecture with independent measures of sleep and circadian traits, we estimated genetic correlations using LD score regression.

CA showed significant positive genetic correlations with self-reported chronotype:

- UKB quantitative chronotype (−2 = definite evening to +2 = definite morning; N = 394,092): $r_g$ = 0.34, $p = 6.27 \times 10^{-14}$

We also estimated genetic correlations between CA and nine accelerometer-derived sleep traits ((Jones, van Hees, et al. 2019; Ferguson et al. 2018); N ≈ 84,000–86,000 across traits) (Fig. 8). Significant negative correlations were observed with sleep timing traits:

- L5 timing: $r_g = -0.36$, $p = 1.30 \times 10^{-7}$
- M10 timing: $r_g = -0.36$, $p = 1.35 \times 10^{-5}$
- Sleep midpoint: $r_g = -0.33$, $p = 6.60 \times 10^{-5}$

No significant genetic correlations were detected with sleep duration, sleep efficiency, number of sleep episodes, or daytime inactivity after correction for multiple testing. A nominal correlation was observed for circadian amplitude ($r_g = 0.20$, $p = 0.013$) and variability in sleep duration ($r_g = -0.31$, $p = 8.1 \times 10^{-3}$), but were not significant after FDR correction.

S6.6 Mendelian randomization

We used two-sample Mendelian randomization (MR) to investigate the bidirectional relationship between CA and chronotype using summary statistics (Fig. S19).

CA → Chronotype

The genetically predicted effect of CA on chronotype was not significant. Using 16 instruments, the IVW random-effects analysis showed no evidence of an effect (β = 0.0111, SE = 0.0182, p = 0.544). Results were consistent across sensitivity methods, with MR-Egger showing no association (β = −0.0090, SE = 0.0607, p = 0.884) and the weighted median estimate similarly null (β = −0.0003, SE = 0.0175, p = 0.986).

Chronotype → CA

In the reverse direction (chronotype as the exposure), MR analyses provided evidence of a strong association with CA across multiple estimators using 94 instruments. IVW random-effects MR indicated a positive effect (β = 0.3278, SE = 0.0565, p = $7.16\times10^{-9}$). Sensitivity analyses were directionally consistent: the weighted median estimate was positive (β = 0.2331, SE = 0.0668, p = $4.85\times10^{-4}$) and MR-Egger regression also suggested a positive association (β = 0.7971, SE = 0.2125, p = $3.09\times10^{-4}$).

Evidence of heterogeneity was observed among SNP-specific causal estimates for the chronotype → CA analysis (IVW Cochran's Q = 164.97, df = 93, p = $6.30\times10^{-6}$; MR-Egger Q = 156.07, df = 92, p = $3.49\times10^{-5}$). The MR-Egger intercept test indicated evidence of directional horizontal pleiotropy (intercept = −0.0116, SE = 0.00506, p = 0.0242). Together, these sensitivity analyses suggest that while the estimated effect was robust in direction across methods, heterogeneity and potential pleiotropy should be considered when interpreting the magnitude of the chronotype → CA effect.

We next examined the effect of genetically predicted CA on the accelerometer-derived activity measures that had a significant genetic correlation with CA: sleep midpoint (MIDP), least-active 5-hour period (L5), and most-active 10-hour period (M10), using 16 genetic instruments (CA COJO, excluding palindromic SNP with intermediate allele frequencies) (Fig. S20). We could not perform the reversed analysis because there were not enough independent variants in the accelerometer GWASs ($n \leq 5$) (Fig. 5B).

CA → Sleep Midpoint (MIDP)

There was no evidence that genetically predicted CA was associated with sleep midpoint. The IVW random-effects estimate was small and non-significant (β = 0.0261, SE = 0.0208, p = 0.208). Sensitivity analyses were consistent with the primary analysis, with no significant associations observed using MR-Egger (β = 0.0336, SE = 0.0694, p = 0.636) or the weighted median method (β = 0.0407, SE = 0.0262, p = 0.121).

CA → L5 (least-active 5-hour period)

Similarly, no association was observed between genetically predicted CA and L5. The IVW estimate was close to null (β = 0.0098, SE = 0.0241, p = 0.684). MR-Egger (β = 0.0518, SE = 0.0796, p = 0.526) and weighted median (β = 0.0305, SE = 0.0279, p = 0.275) analyses were directionally consistent and non-significant.

CA → M10 (most-active 10-hour period)

For M10, IVW analysis again showed no evidence of an effect (β = 0.0174, SE = 0.0162, p = 0.284). MR-Egger (β = 0.0684, SE = 0.0586, p = 0.262) and weighted median (β = 0.0117, SE = 0.0247, p = 0.637) estimates were likewise non-significant.

Across all three accelerometer-derived activity measures with significant negative genetic correlations with CA (MIDP, L5, and M10), there was no evidence to support a causal effect of genetically predicted CA.

S6.6. Association between circadian acceleration PRS and chronotype

We tested whether the polygenic score for circadian acceleration (CA-PRS) was associated with self-reported chronotype in the full UKB samples excluding the proteomics subset. Using linear regression with "definitely morning" as the reference category, higher CA-PRS was progressively associated with later chronotype categories. Compared with definite morning types, individuals reporting "rather morning" showed a small decrease in CA-PRS ($\beta = -0.016$, $p = 2.6 \times 10^{-4}$), while "Don't know" chronotypes showed a similar but weaker shift ($\beta = -0.012$, $p = 0.04$). Stronger associations were observed for evening chronotypes: "rather evening" individuals had substantially lower CA-PRS values ($\beta = -0.030$, $p = 4.1 \times 10^{-11}$), and "definitely evening" individuals showed the largest difference relative to morning types ($\beta = -0.043$, $p = 5.7 \times 10^{-11}$).

The sample comprised 380,075 individuals with chronotype information, including 90,804 "definitely morning", 123,098 "rather morning", 39,487 "don't know", 96,875 "rather evening", and 29,811 "definitely evening" participants. Overall, these results indicate a graded relationship between genetic predisposition to circadian acceleration and chronotype.

S7. Ethics statement

Study subjects in FinnGen provided informed consent for biobank research, based on the Finnish Biobank Act. Alternatively, separate research cohorts, collected prior the Finnish Biobank Act came into effect (in September 2013) and start of FinnGen (August 2017), were collected based on study-specific consents and later transferred to the Finnish biobanks after approval by Fimea (Finnish Medicines Agency), the National Supervisory Authority for Welfare and Health. Recruitment protocols followed the biobank protocols approved by Fimea. The Coordinating Ethics Committee of the Hospital District of Helsinki and Uusimaa (HUS) statement number for the FinnGen study is Nr HUS/990/2017.

The FinnGen study is approved by Finnish Institute for Health and Welfare (permit numbers: THL/2031/6.02.00/2017, THL/1101/5.05.00/2017, THL/341/6.02.00/2018, THL/2222/6.02.00/2018, THL/283/6.02.00/2019, THL/1721/5.05.00/2019 and THL/1524/5.05.00/2020), Digital and population data service agency (permit numbers: VRK43431/2017-3, VRK/6909/2018-3, VRK/4415/2019-3), the Social Insurance Institution (permit numbers: KELA 58/522/2017, KELA 131/522/2018, KELA 70/522/2019, KELA 98/522/2019, KELA 134/522/2019, KELA 138/522/2019, KELA 2/522/2020, KELA 16/522/2020), Findata permit numbers THL/2364/14.02/2020, THL/4055/14.06.00/2020, THL/3433/14.06.00/2020, THL/4432/14.06/2020, THL/5189/14.06/2020, THL/5894/14.06.00/2020, THL/6619/14.06.00/2020, THL/209/14.06.00/2021, THL/688/14.06.00/2021, THL/1284/14.06.00/2021, THL/1965/14.06.00/2021, THL/5546/14.02.00/2020, THL/2658/14.06.00/2021, THL/4235/14.06.00/2021, Statistics Finland (permit numbers: TK-53-1041-17 and TK/143/07.03.00/2020 (earlier TK-53-90-20) TK/1735/07.03.00/2021, TK/3112/07.03.00/2021) and Finnish Registry for Kidney Diseases permission/extract from the meeting minutes on 4th July 2019.

The Biobank Access Decisions for FinnGen samples and data utilized in FinnGen Data Freeze 11 include: THL Biobank BB2017_55, BB2017_111, BB2018_19, BB_2018_34, BB_2018_67, BB2018_71, BB2019_7, BB2019_8, BB2019_26, BB2020_1, BB2021_65, Finnish Red Cross Blood Service Biobank 7.12.2017, Helsinki Biobank HUS/359/2017, HUS/248/2020, HUS/430/2021 §28, §29, HUS/150/2022 §12, §13, §14, §15, §16, §17, §18, §23, §58, §59, HUS/128/2023 §18, Auria Biobank AB17-5154 and amendment #1 (August 17 2020) and amendments BB_2021-0140, BB_2021-0156 (August 26 2021, Feb 2 2022), BB_2021-0169, BB_2021-0179, BB_2021-0161, AB20-5926 and amendment #1 (April 23 2020) and it´s modifications (Sep 22 2021), BB_2022-0262, BB_2022-0256, Biobank Borealis of Northern Finland_2017_1013, 2021_5010, 2021_5010 Amendment, 2021_5018, 2021_5018 Amendment, 2021_5015, 2021_5015 Amendment, 2021_5015 Amendment_2, 2021_5023, 2021_5023 Amendment, 2021_5023 Amendment_2, 2021_5017, 2021_5017 Amendment, 2022_6001, 2022_6001 Amendment, 2022_6006 Amendment, 2022_6006 Amendment, 2022_6006 Amendment_2, BB22-0067, 2022_0262, 2022_0262 Amendment, Biobank of Eastern Finland 1186/2018 and amendment 22§/2020, 53§/2021, 13§/2022, 14§/2022, 15§/2022, 27§/2022, 28§/2022, 29§/2022, 33§/2022, 35§/2022, 36§/2022, 37§/2022, 39§/2022, 7§/2023, 32§/2023, 33§/2023, 34§/2023, 35§/2023, 36§/2023, 37§/2023, 38§/2023, 39§/2023, 40§/2023, 41§/2023, Finnish Clinical Biobank Tampere MH0004 and amendments (21.02.2020 & 06.10.2020), BB2021-0140 8§/2021, 9§/2021, §9/2022, §10/2022, §12/2022, 13§/2022, §20/2022, §21/2022, §22/2022, §23/2022, 28§/2022, 29§/2022, 30§/2022, 31§/2022, 32§/2022, 38§/2022, 40§/2022, 42§/2022, 1§/2023, Central Finland Biobank 1-2017, BB_2021-0161, BB_2021-0169, BB_2021-0179, BB_2021-0170, BB_2022-0256, BB_2022-0262, BB22-0067, Decision allowing to continue data processing until 31st Aug 2024 for projects: BB_2021-0179, BB22-0067,BB_2022-0262, BB_2021-0170, BB_2021-0164, BB_2021-0161, and BB_2021-0169, and Terveystalo Biobank STB 2018001 and amendment 25th Aug 2020, Finnish Hematological Registry and Clinical Biobank decision 18th June 2021, Arctic biobank P0844: ARC_2021_1001.

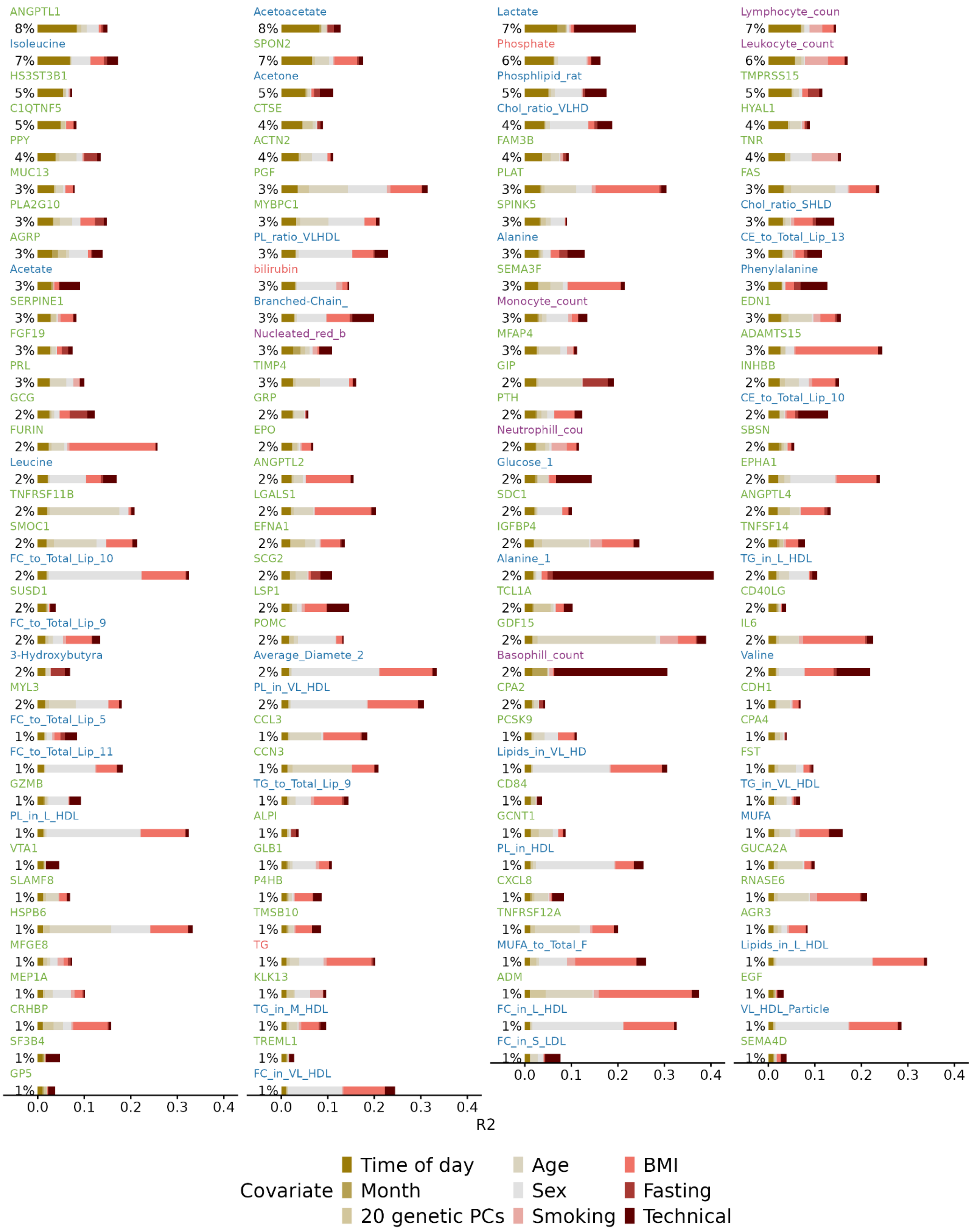


**Fig. S1. Proportion of biomarker variance explained by covariates.** Top 134 biomarkers with time-of-day $R^2 > 1\%$. Biochemistry biomarkers labels are shown in orange, cell counts in purple, metabolites in blue, and proteins in green. Grouped technical covariates were: assessment centre and platform-specific:

Olink proteomics (disorder selected, batch, panel date), NMR-metabolomics (batch, spectrometer, time analysis), biochemistry (dilution factor, acquisition time) cell counts (device id, acquisition time).

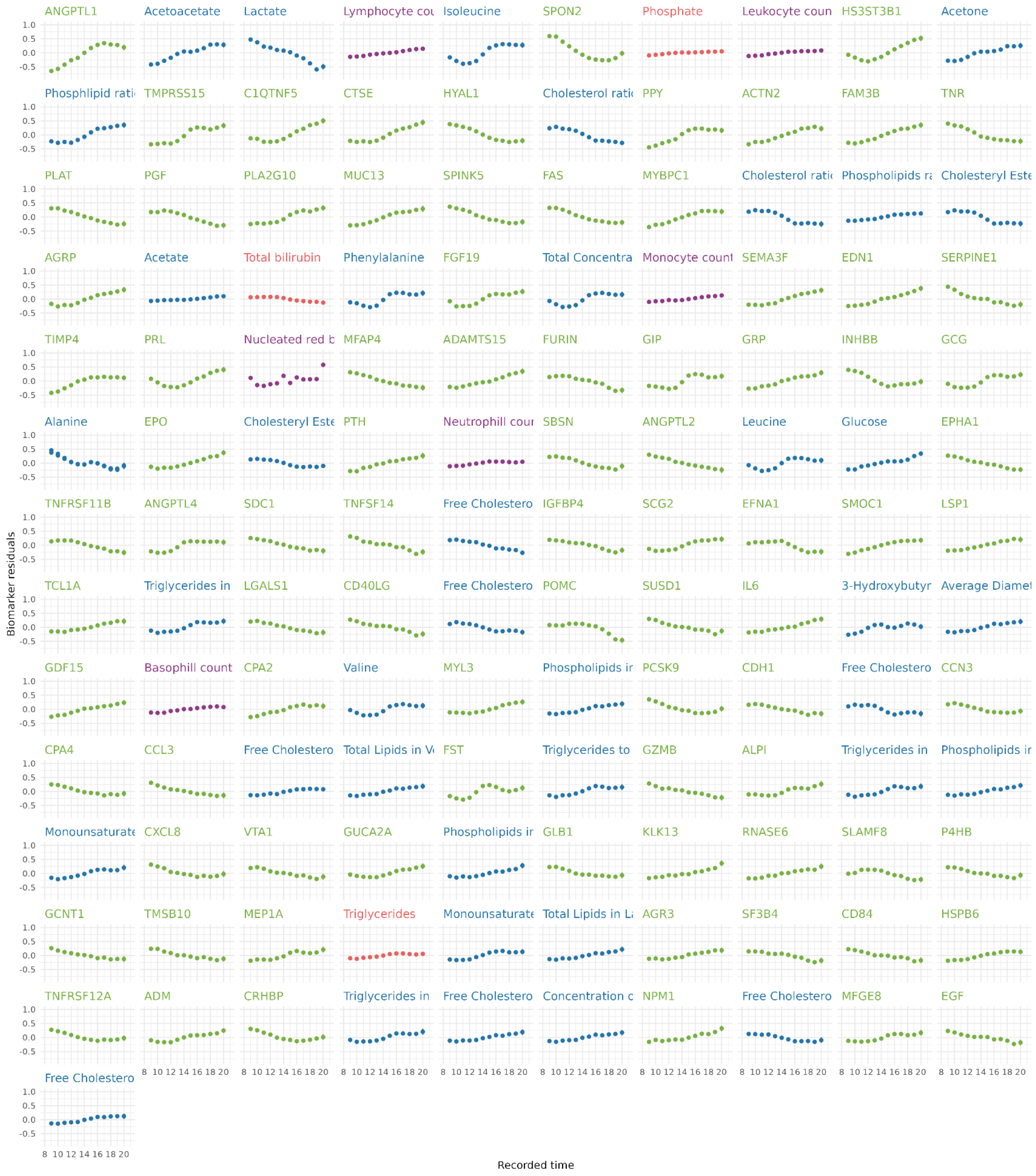


**Fig. S2. Mean hourly values of 134 biomarkers with R2 time-of-day > 1%.** Hourly mean values (y-axis) in the interval 9h to 20h (x-axis). Vertical bars show 95% CIs. Biochemistry biomarkers labels are shown in orange, cell counts in purple, metabolites in blue, and proteins in green. The biomarker values are the residuals from a linear regression model with their values as outcome and sex, age, fasting hours, month of assessment and all technical covariates described in Methods as covariates.

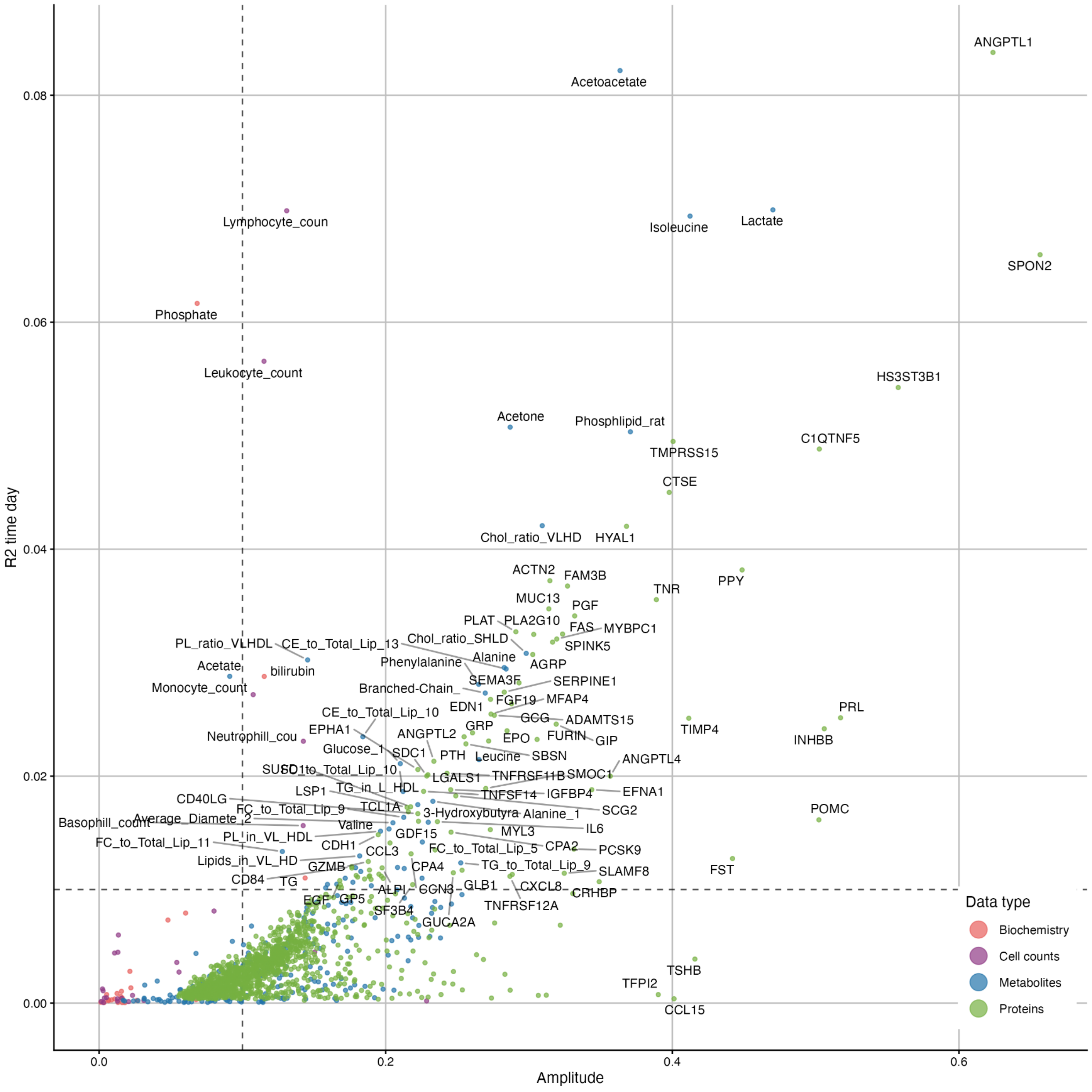


**Fig. S3. Relationship between amplitude and time-of-day variance explained for 1,545 biomarkers.** Labels are shown for all biomarkers with both amplitude > 0.1 and time of day R2 > 0.01, including extreme values with R2 > 0.02 or amplitude ~0.4. Low amplitude but high variance explained are for cell count/biochemistry measures where sample sizes are 9-fold of those for proteins, so reflect higher power (reduced error on estimates).

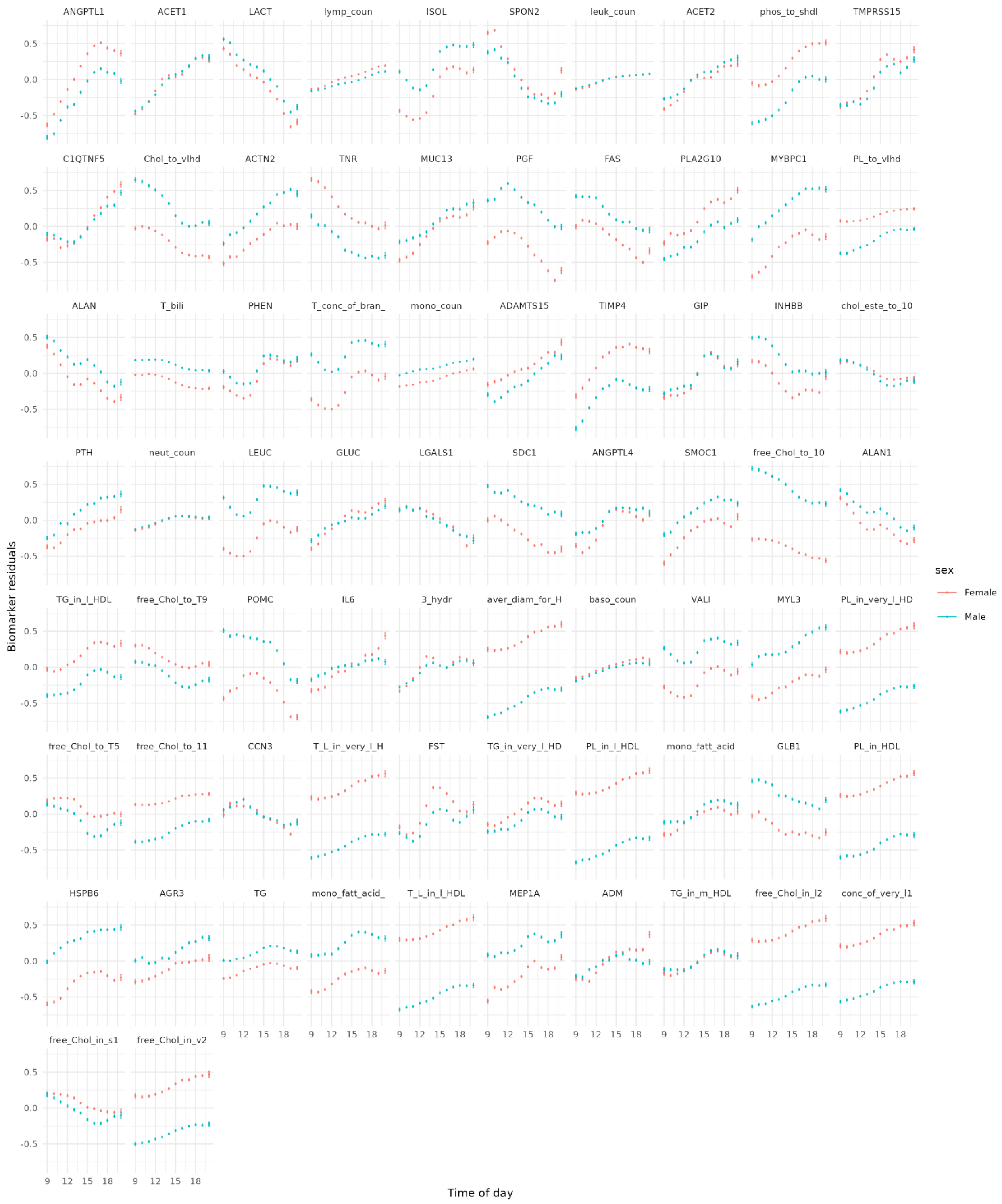


**Fig. S4. Mean hourly values of 72 biomarkers with R2 time-of-day > 1% and significant time x sex interaction.** Hourly mean values (y-axis) in the interval 9h to 20h (x-axis). Vertical bars show 95% CIs in SDU. The biomarker values are the residuals from a linear regression model with (normalised) biomarker values as outcome and all technical covariates described in Methods as covariates.

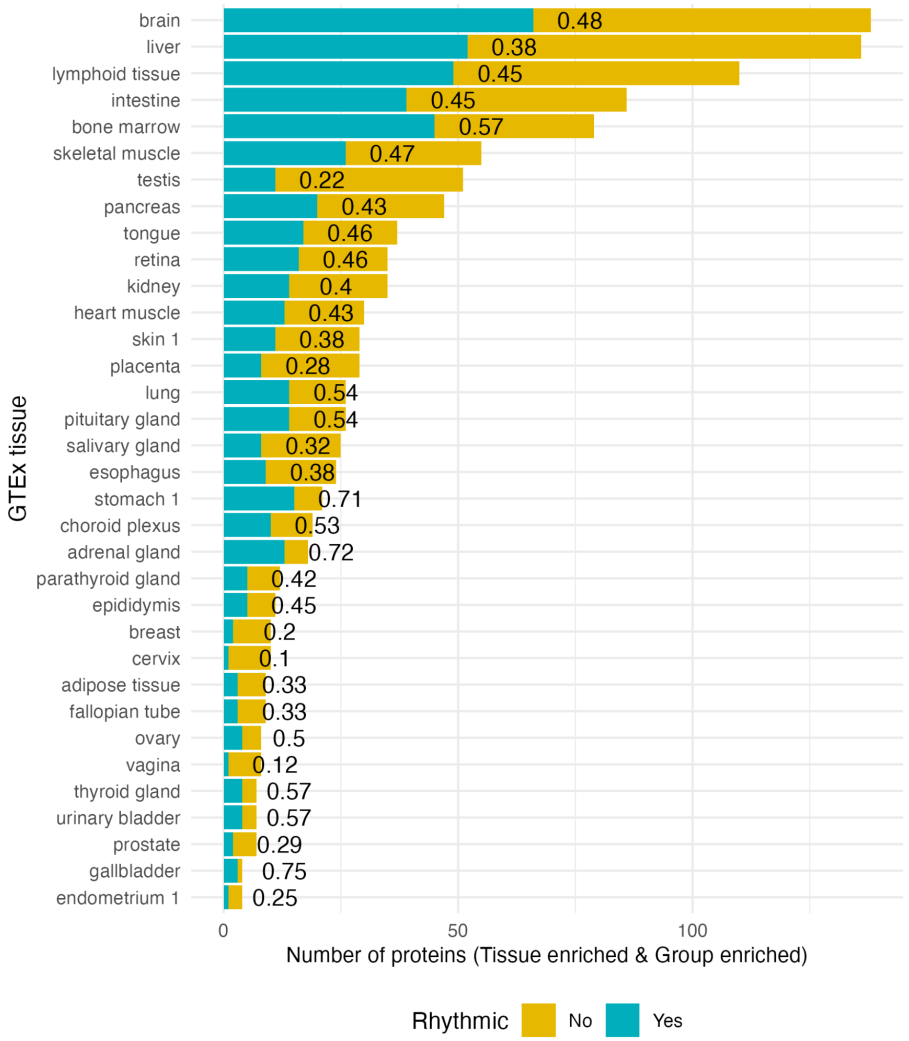


**Fig. S5. Number of 4-fold expression-enriched proteins per tissue.** The plot shows protein counts (x-axis) for each of the 34 GTEx tissues expressing at least 1 *Tissue enriched or Group enriched* rhythmic protein (y-axis). Proteins labelled as 'Rhythmic' had a significant effect of time in a harmonic model, after multiple testing correction. There were 746 unique UniProt IDs in all the categories, where 323 were labelled as rhythmic.

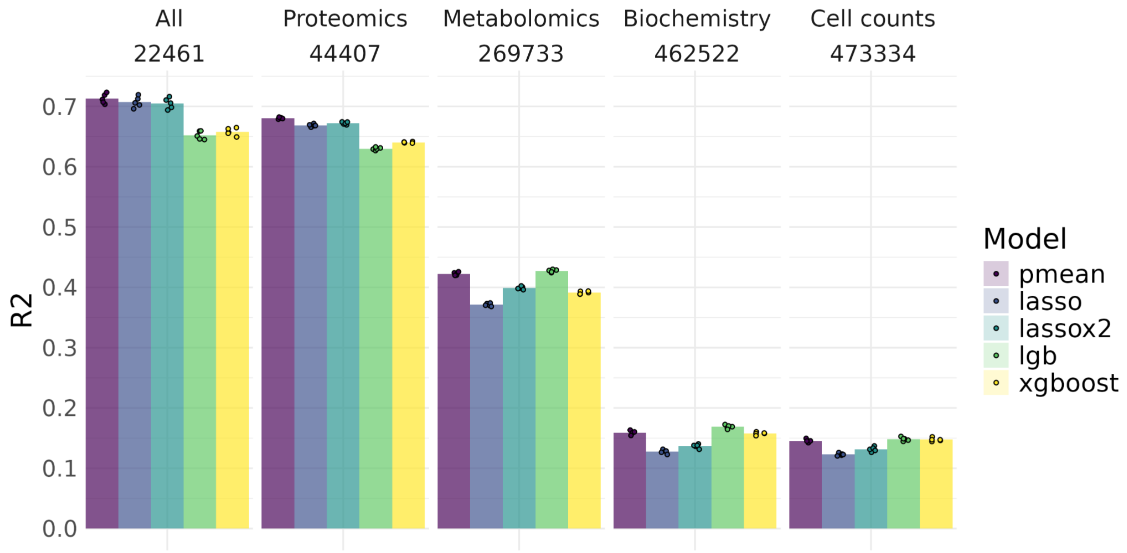


**Fig. S6. Time-of-day prediction model benchmark.** Comparison in prediction accuracy R2 between the predicted values and the recorded time-of-day of the blood sample. Each data point represents the R2 from a different 5-fold cross validation split (80% training, 20% testing). The results are shown by training data type (All, proteomics, metabolomics, biochemistry and cell counts, sample size below label). The pmean model was built by taking the mean prediction of all four models.

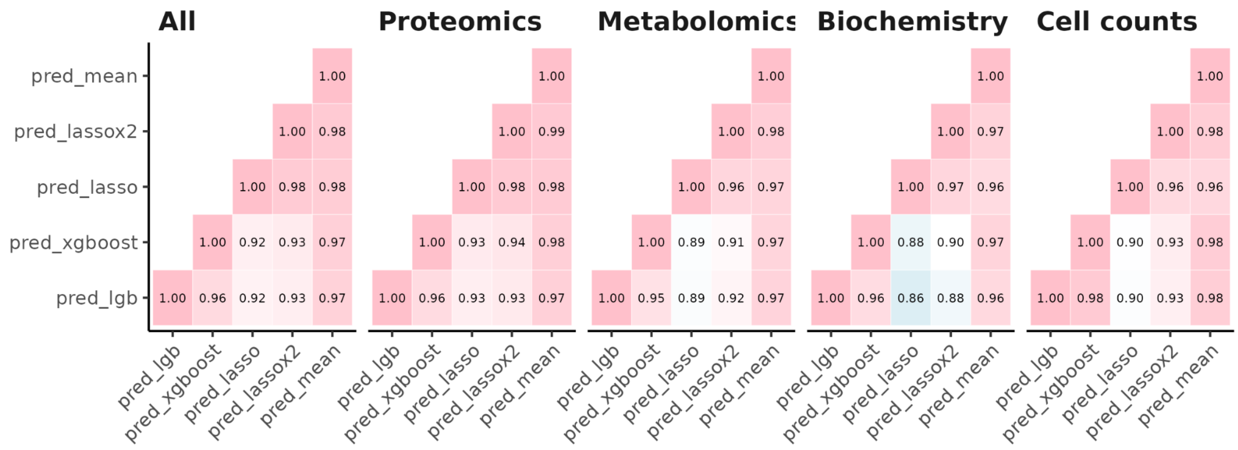


**Fig. S7. Correlation estimates between prediction models within biomarker data type.** For each data type, the Pearson correlation estimates reflect the 4 different models: LASSO, LASSO_x2, XGBOOST and LIGHTGB and their mean (pred_mean).

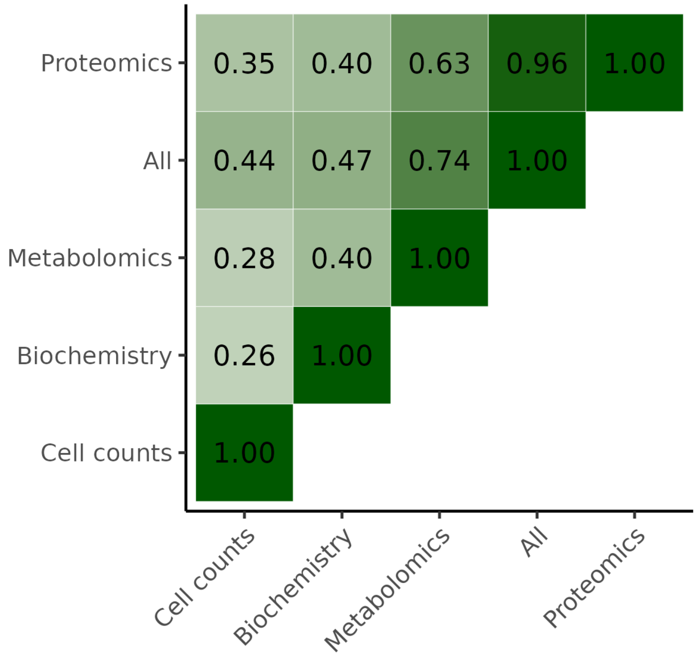


**Fig. S8. Correlation estimates between the mean time-of-day prediction (4 models) across biomarker data types.** For each data platform, the predicted values were the mean prediction across 4 models: LASSO, LASSO_x2, XGBOOST and LIGHTGB.

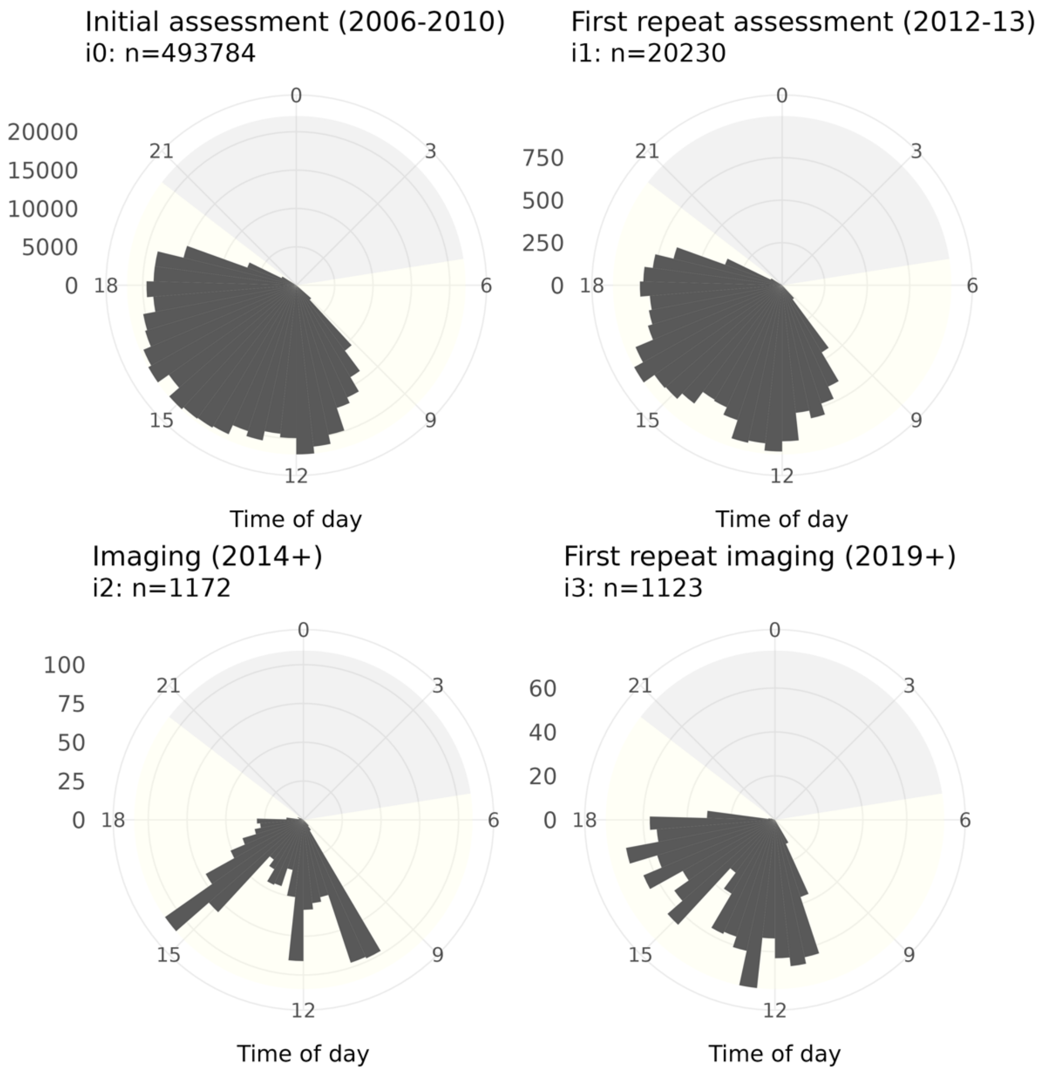


**Fig. S9. Histograms of time-of-day for UKB instances with blood biomarkers.**

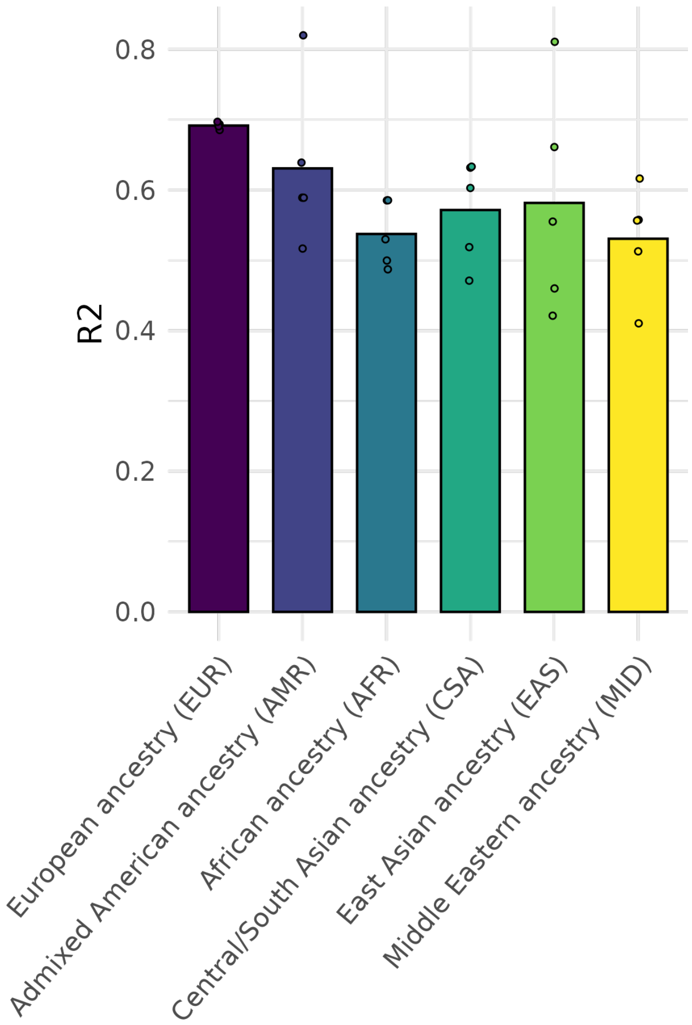


**Fig. S10. Time-of-day prediction accuracy transferability across genetically-inferred ancestries.** Comparison in prediction accuracy R2 between the predicted values and the recorded time-of-day of the blood sample, where the models were trained on the full 44,407 individuals. Each data point represents the R2 from a different 5-fold cross validation split (80% training on all, 20% testing on each ancestry group separately), with the bar showing the mean R2.

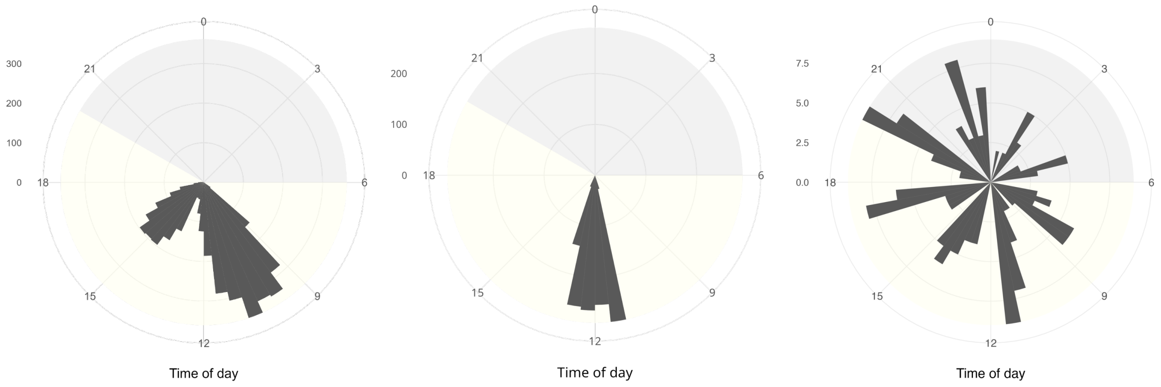


**Fig. S11. Histograms of recorded time-of-day of blood sampling for CKB (left), FinnGen (middle) and TREASURE (right).**

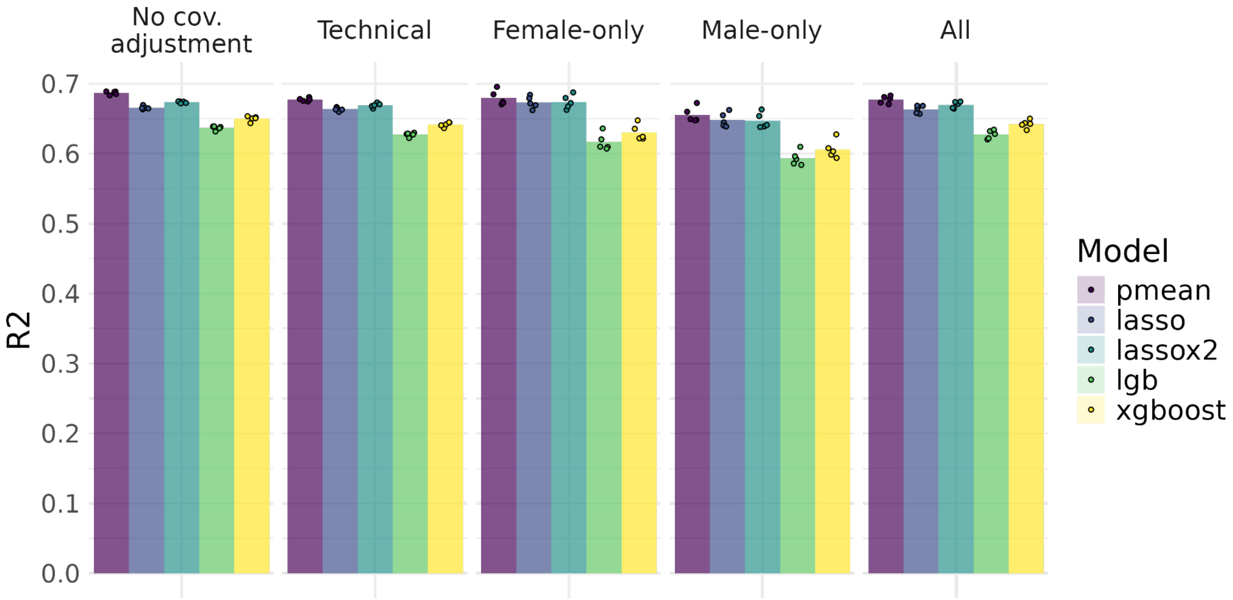


**Fig. S12. Time-of-day prediction model benchmark across different biomarker covariate adjustments.** Comparison in prediction accuracy R2 between the predicted values and the recorded time-of-day of the blood sample. Models were run with different covariate adjustments: no covariate adjustments, only technical covariates for both sexes, sex-specfic training on only technical covariates and all covariates (technical, age, sex, PCs, fasting h, month of attending). Each data point represents the R2 from a different 5-fold cross validation split (80% training, 20% testing). The pmean model was built by taking the mean prediction of all other models.

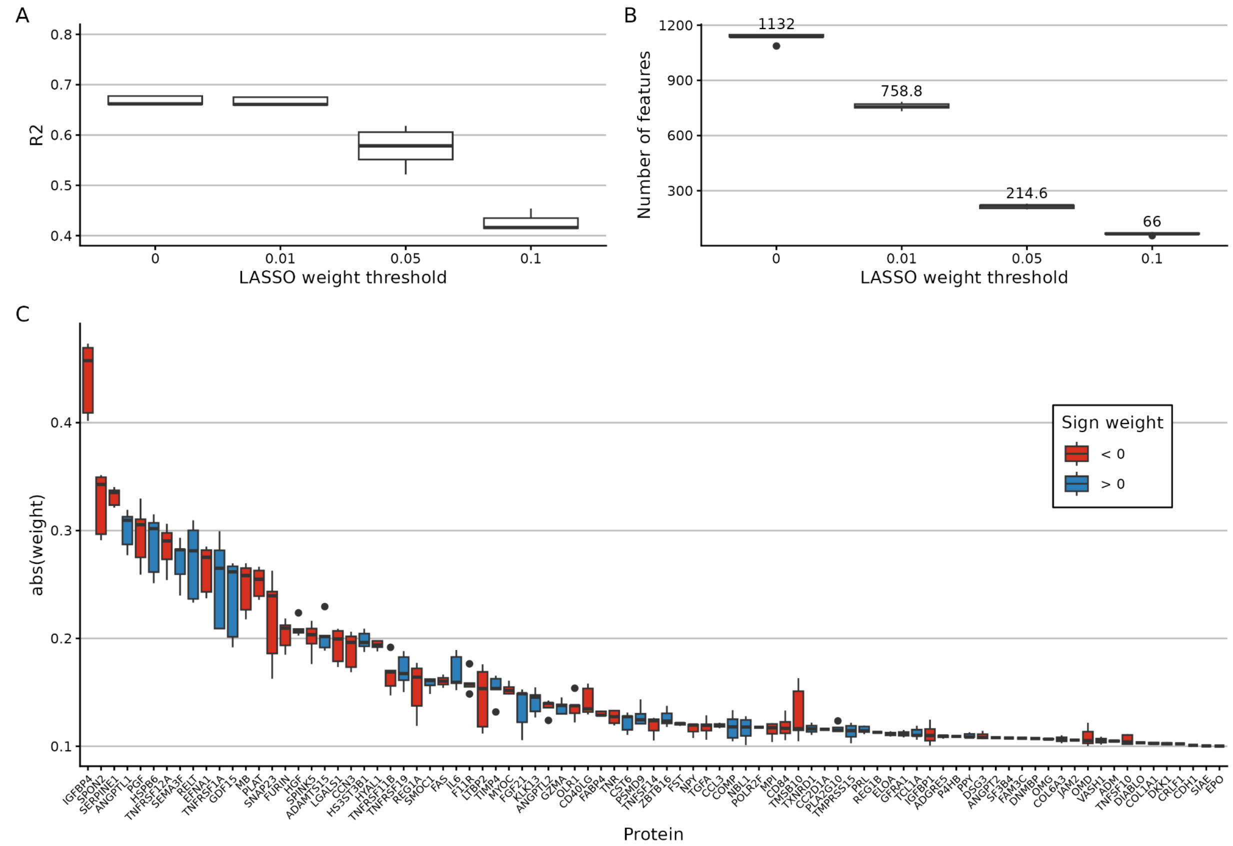


**Fig. S13. LASSO feature selection. A.** Time-of-day prediction accuracy (R2) for the proteomic models with 1,459 features for the training 5-fold cross-validation sets, across four weight thresholds. **B.** Number of features with weights different from 0 across four thresholds. **C.** Feature weights for the 66 proteins with absolute weights > 0.1, across the 5-fold cross validation sets. Color indicates the sign of the LASSO weight.

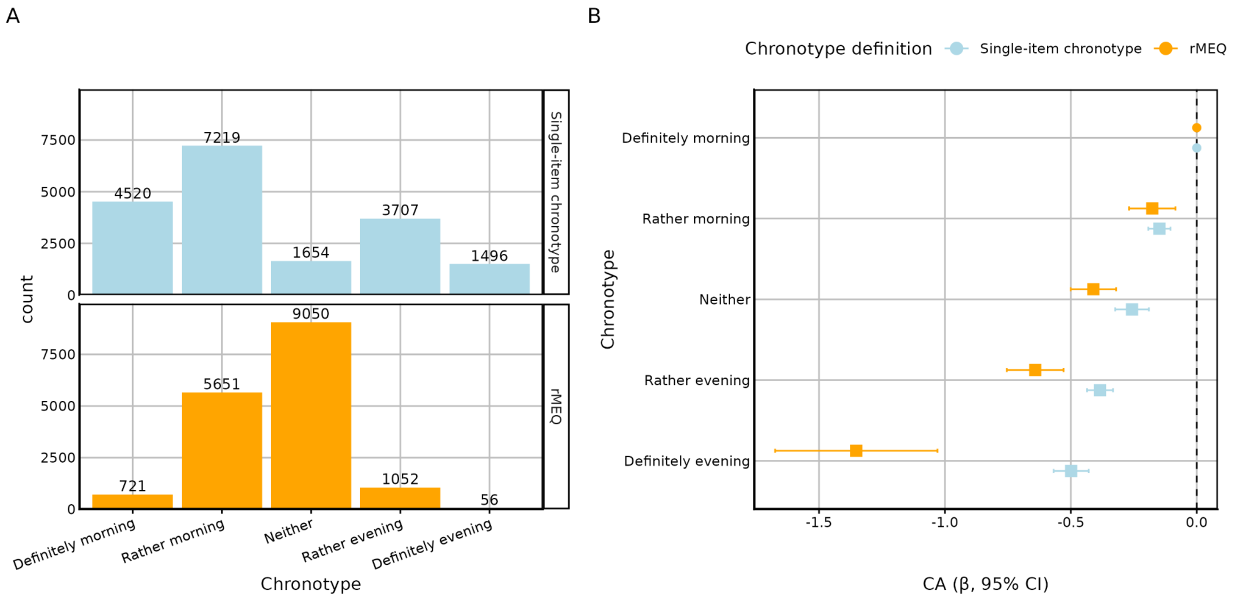


**Fig. S14. Comparison between chronotype definitions and their association with blood circadian acceleration (CA). A.** Distribution of categorial chronotypes. Chronotype categories were obtained from the UKB follow-up sleep questionnaire, with "Single-item chronotype" corresponding to the question "One hears about 'morning-types' and 'evening-types.' Which one of these types do you consider yourself to be?" and the rMEQ (reduced Morningness-Eveningness Questionnaire) chronotype is based on the combined score across 5 sleep-related questions(Adan and Almirall 1991). B. Chronotype associations with blood CA. All CA effect sizes come from linear regression models, adjusted for sex, age, assessment centre and the first 20 genetic principal components. Beta estimates and 95% CI are shown in the y axis in hours. The reference level (Definitely morning) is indicated by a dot, while associations passing an FDR statistical significance threshold of 5% are shown in bold colors.

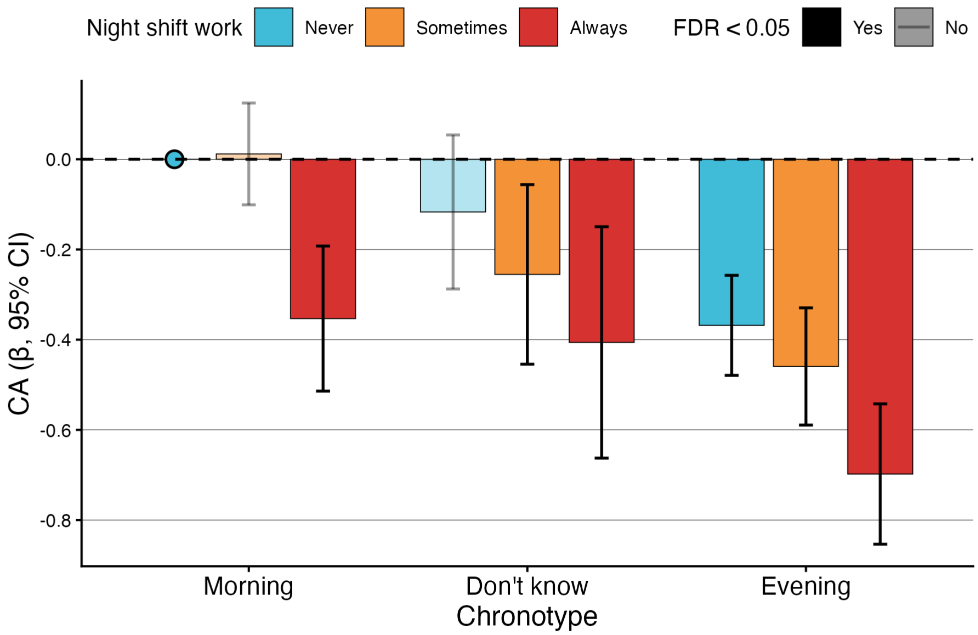


**Fig. S15. Chronotype x Night shift work associations with blood circadian acceleration (CA).** All CA effect sizes come from linear regression models, adjusted for sex, age, assessment centre and the first 20 genetic principal components. Beta estimates and 95% CI are shown in the y axis in hours. The reference level (Morning-Alwaws night shifts) is indicated by a blue dot, while associations passing an FDR statistical significance threshold of 5% are shown in bold colors.

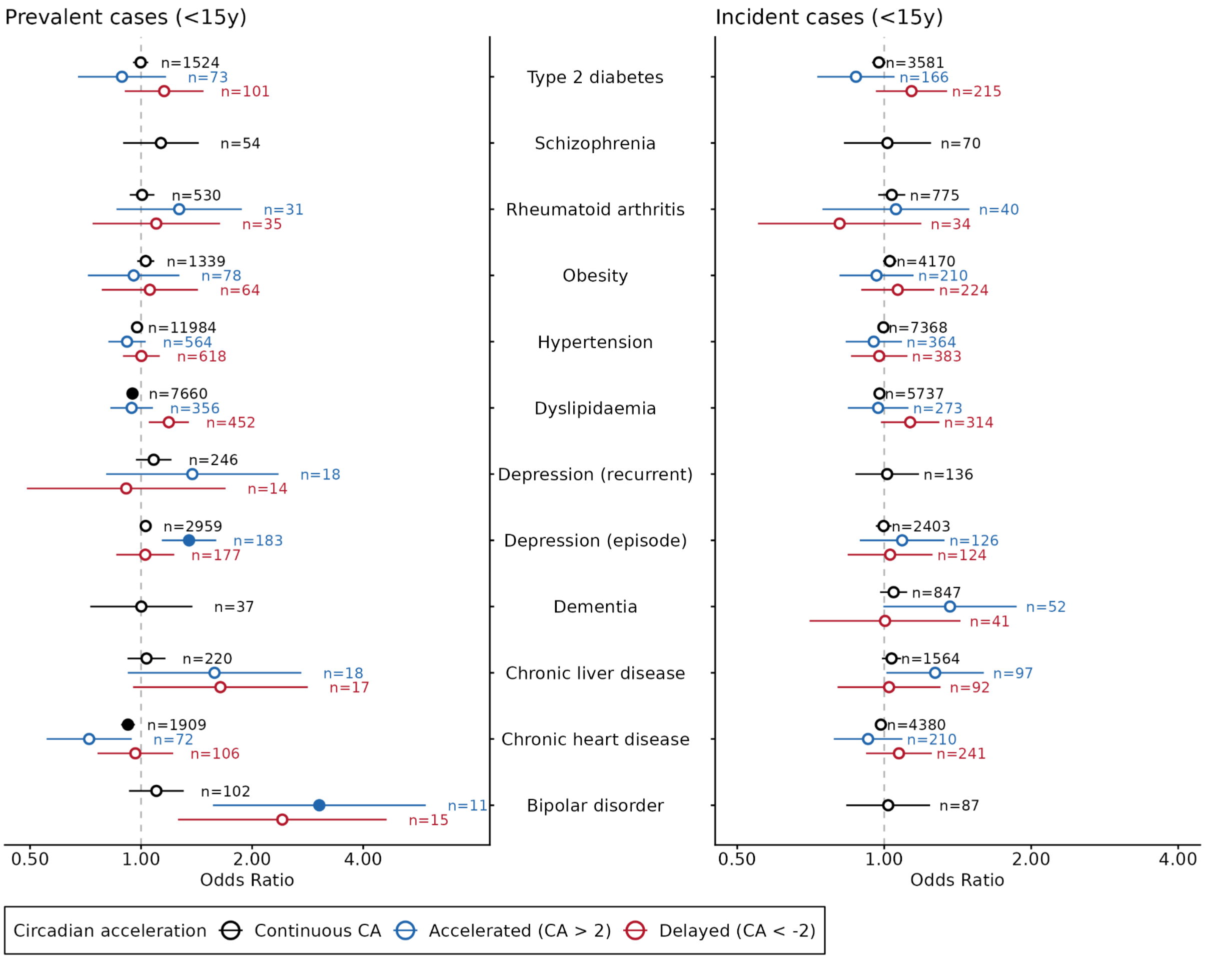


**Fig. S16. Blood circadian acceleration (CA) associations with disease status in the UKB.** Odds ratios and 95% CI (x-axis) of CA levels for models including prevalent cases (left) and incident cases (right) of 12 diseases (y-axis) with hypothesized circadian involvement. For each disease and timeline, we show the total number of cases in black, the number of accelerated cases with CA > 2 in blue and the number of delayed cases with CA < -2 in red. Disease groups with <10 cases were excluded from analysis. CA risk was evaluated in three models: as a continuous variable (black) and as a categorial variable comparing both accelerated (blue) and delayed (red) to middle CA (-2 > CA < 2). We included disease cases for a 30 year period, split in 15 years before the blood sampling took place (prevalent cases) and 15 years after the blood sampling (incident cases). All CA odds ratios come from logistic regression models, adjusted for sex, age, assessment centre, chronotype, season of blood sampling, Townend Deprivation Index, BMI, smoking status and the first 20 genetic principal components. Associations passing an FDR statistical significance threshold of 5% are shown as a filled dot.

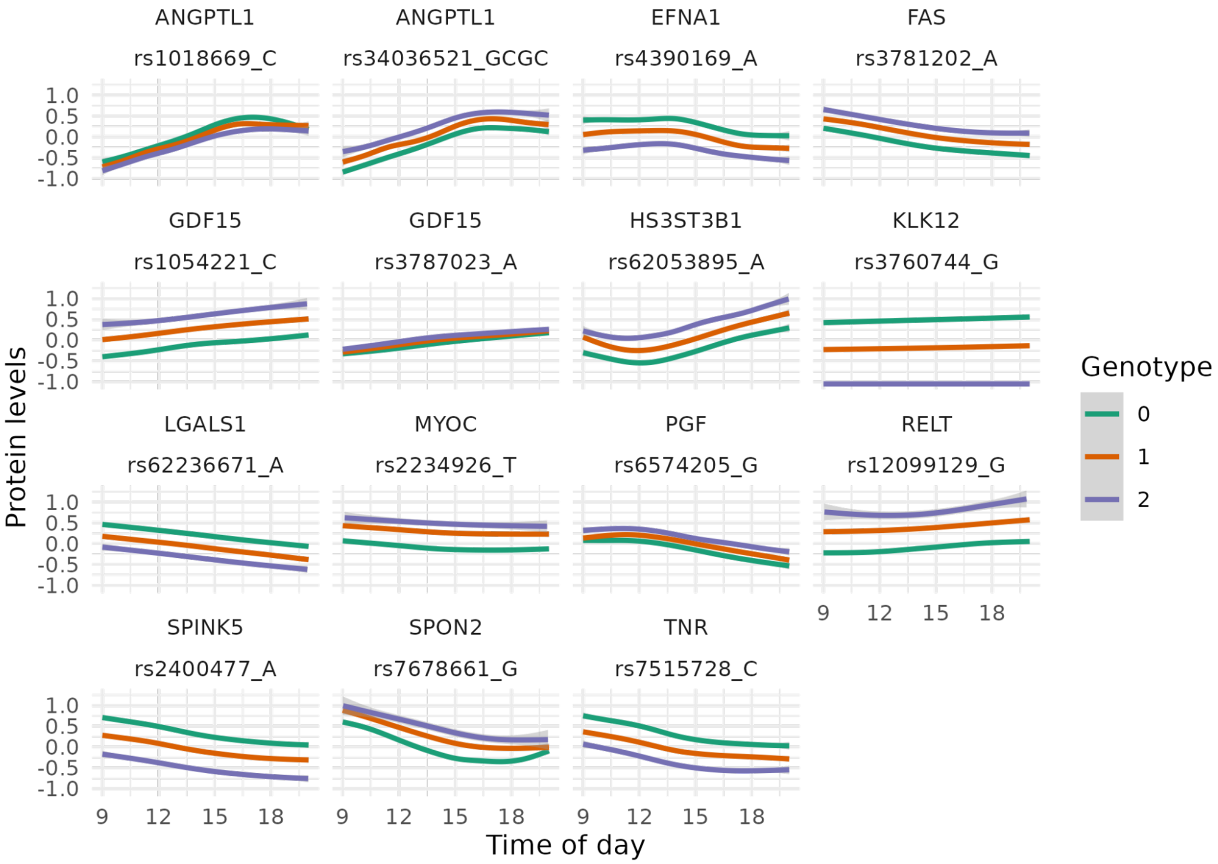


**Fig. S17. Relationship between time of day and protein levels for 15 CA pQTL genotypes.** The protein values (y-axis) are shown for each available time of blood sampling (x-axis). For each SNP-Olink protein-pair, the SNP rsid is shown together with the alternative allele, for which the color represents the number of copies.

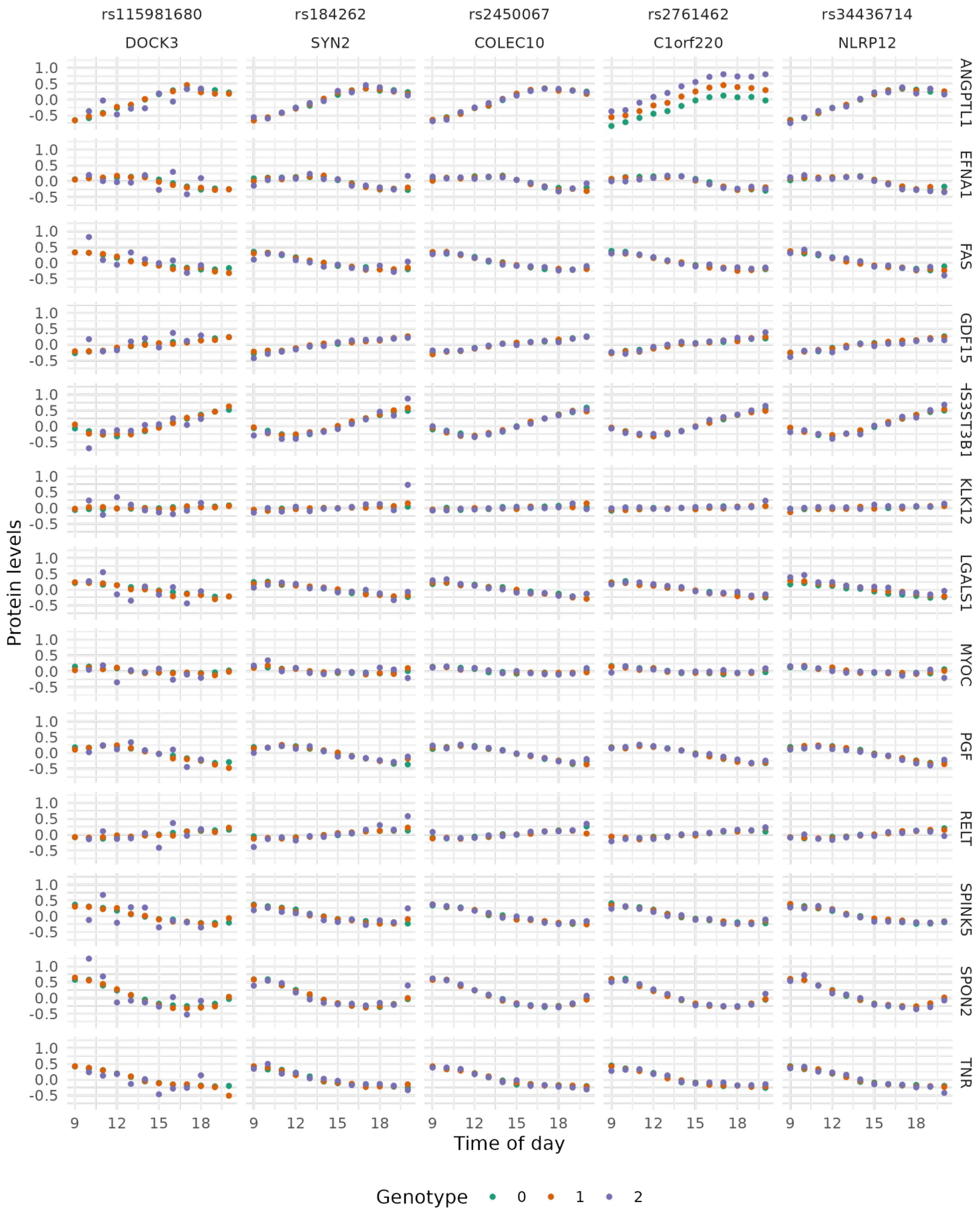


**Fig. S18. Relationship between time of day and protein levels for 5 CA pQTL genotypes.** The hourly mean protein values (y-axis) are shown for each available time of blood sampling (x-axis).

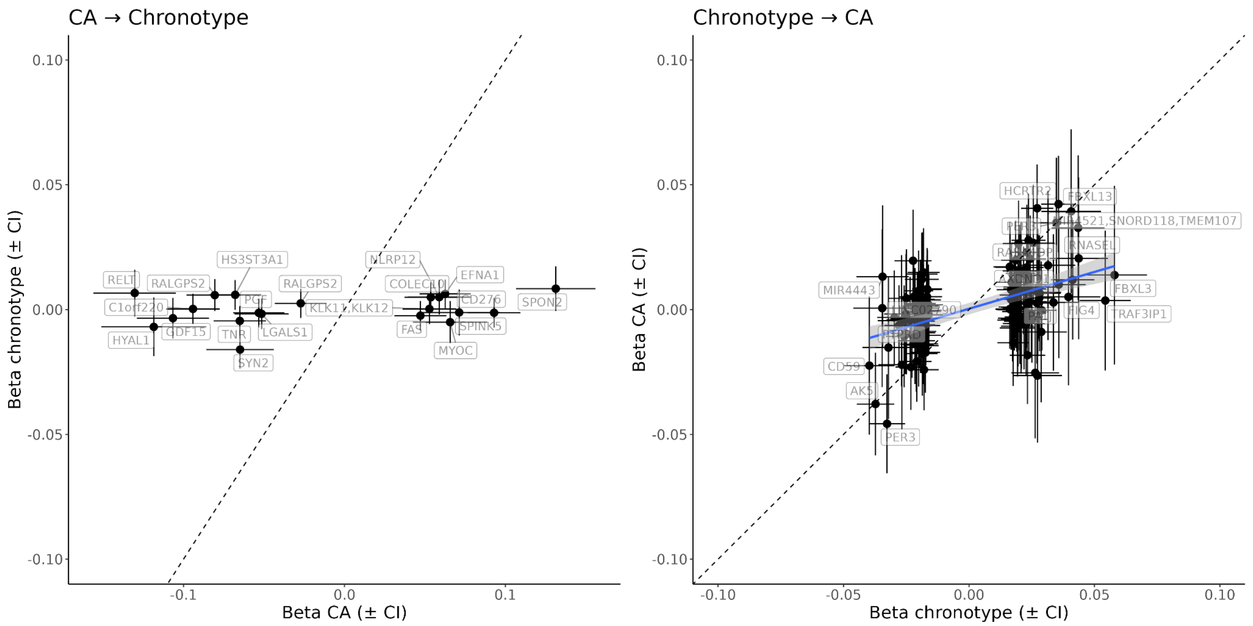


**Fig. S19. GWAS effect size comparison.** Relationship between effect sizes from top CA GWAS (17 COJO SNPs) and chronotype GWAS (left) and top chronotype GWAS (94 COJO SNPs) and CA GWAS (right). The dashed line shows diagonal. Mendelian Randomization suggested a causal relationship between chronotype → CA, with the regression line shown in the plot in blue. Labels are shown for the closest genes (<10kb window).

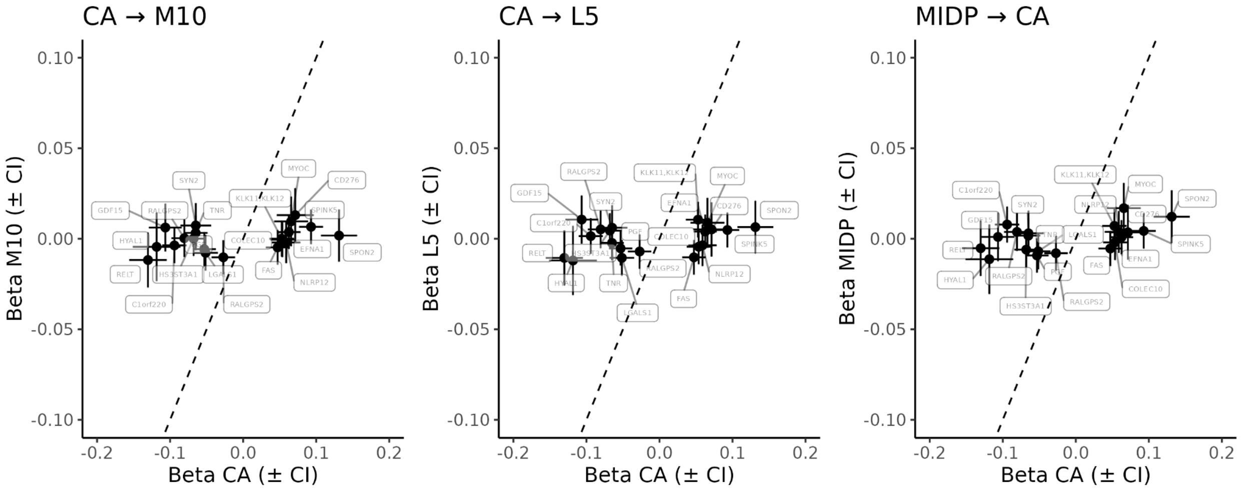


**Fig. S20. GWAS effect size comparison for CA and** accelerometry-derived traits**.** Relationship between effect sizes from top CA GWAS (17 COJO SNPs) and significant genetic correlation accelerometry-derived traits (M10, L5 and MIDP). No significant results were established using Mendelian Randomization. Labels are shown for the closest genes (<10kb window). The dashed line shows diagonal.

**Table S1.**

| Data type | UKB Category ID | Number of variables | Instance 0 | Instance 1 | Instance 2 | Instance 3 |
|---|---|---|---|---|---|---|
| Blood biochemistry | 17518 | 30 | 469,075 | 17,856 | | |
| Blood cell counts | 100081 | 31 | 476,952 | 19,392 | 5,853 | |
| NMR metabolomics (Nightingale) | 220 | 251 | 269,166 | 16,665 | | |
| Proteomics (Olink) | 1838 | 2,924/ | 53,014 | | 1,172 | 1,123 |

Available UK Biobank blood measurements.

**Table S2.**

| | UKB (n = 51701) | CKB (n = 3965) | FinnGen (n = 1265) | TREASURE (n = 12) |
|---|---|---|---|---|
| **Age** | | | | |
| Mean | 56.8 (8.2) | 57.4 (11.7) | 49.0 (13.9, n=1199) | 24.1 (3.0) |
| Range | 39 - 70 | 30.2 - 78.5 | 18-71 (n=1199) | 19 - 28 |
| **Sex** | | | | |
| Female | 27928 (54.0%) | 2134 (53.8%) | 620 (51.7%, n=1199) | 4 (33.3%) |
| Male | 23773 (46.0%) | 1831 (46.2%) | 579 (48.3%, n=1199) | 8 (66.7%) |
| **Ethnicity** | | | | |
| European ancestry (EUR) | 44529 (86.1%) | - | 1265 (100%) | 12 (100%) |
| Admixed American ancestry (AMR) | 97 (0.2%) | - | - | - |
| African ancestry (AFR) | 888 (1.7%) | - | - | - |
| Central/South Asian ancestry (CSA) | 902 (1.7%) | - | - | - |
| East Asian ancestry (EAS) | 256 (0.5%) | 3965 (100%) | - | - |
| Middle Eastern ancestry (MID) | 297 (0.6%) | - | - | - |
| **BMI** | | | | |
| Mean | 27.5 (4.8) | 23.9 (3.64) | 26.5 (4.4) (n=93) | 23.5 (1.5) |
| Range | 14.3 - 69 | 12.9 - 41.4 | 18-53.7 (n=93) | 21-25.9 |
| **Smoking** | | | | |
| Never | 27967 (54.1%) | 2267 (57.2%) | 650 (56.4%, n=1152) | - |
| Previous | 18039 (34.9%) | 302 (7.6%) | 383 (33.3%, n=1152) | - |

| | | | | |
|---|---|---|---|---|
| Current | 5447 (10.5%) | Current regular: 1193 (30.1%)<br>Current occasional: 203 (5.1%) | 119 (10.3%, n=1152) | - |

Cohort demographics.

**Data S1. (separate file)**
Variance explained by blood biomarker covariates.

**Data S2. (separate file)**
Harmonic parameter estimates for blood biomarkers.

**Data S3. (separate file)**
Summary of phenotypic data in proteomic UKB subset.

**Data S4. (separate file)**
Summary of phenotypic data in proteomic UKB subset within extreme CA categories (Accelerated: CA > 2 & Delayed (CA < -2).

**Data S5. (separate file)**
Independent subset of variants in CA GWAS from COJO, including nearest-gene annotations from ANNOVAR.

**Data S6. (separate file)**
Polyfun/FINEMAP results for genome-wide analysis of CA GWAS.

**Data S7. (separate file)**
Genetic correlations between CA, self-reported chronotype and accelerometry-derived sleep measures.

**Data S8. (separate file)**
Genetic correlations between CA and selected diseases and traits.

**Data S9 (separate file)**
FinnGen Consortium author list.

"ANNOVAR: Functional Annotation of Genetic Variants from next-Generation Sequencing Data Nucleic." 2010. *Nucleic Acids Research* 38.

Aragam, Krishna G., Tao Jiang, Anuj Goel, et al. 2022. "Discovery and Systematic Characterization of Risk Variants and Genes for Coronary Artery Disease in over a Million Participants." *Nature Genetics* 54 (12): 1803–1815.

Astle, William J., Heather Elding, Tao Jiang, et al. 2016. "The Allelic Landscape of Human Blood Cell Trait Variation and Links to Common Complex Disease." *Cell* 167 (5): 1415–1429.e19.

Benner, Christian, Chris C. A. Spencer, Aki S. Havulinna, Veikko Salomaa, Samuli Ripatti, and Matti Pirinen. 2016. "FINEMAP: Efficient Variable Selection Using Summary Data from Genome-Wide Association Studies." *Bioinformatics (Oxford, England)* 32 (10): 1493–1501.

Bulik-Sullivan, Brendan K., Po-Ru Loh, Hilary K. Finucane, et al. 2015. "LD Score Regression Distinguishes Confounding from Polygenicity in Genome-Wide Association Studies." *Nature Genetics* 47 (3): 291–295.

Chen, Tianqi, and Carlos Guestrin. 2016. "XGBoost: A Scalable Tree Boosting System." *Proceedings of the 22nd ACM SIGKDD International Conference on Knowledge Discovery and Data Mining* (New York, NY, USA), KDD '16, August 13, 785–794.

Chen, Zhengming, Junshi Chen, Rory Collins, et al. 2011. "China Kadoorie Biobank of 0.5 Million People: Survey Methods, Baseline Characteristics and Long-Term Follow-Up." *International Journal of Epidemiology* 40 (6): 1652–1666.

Christakoudi, Sofia, Evangelos Evangelou, Elio Riboli, and Konstantinos K. Tsilidis. 2021. "GWAS of Allometric Body-Shape Indices in UK Biobank Identifies Loci Suggesting Associations with Morphogenesis, Organogenesis, Adrenal Cell Renewal and Cancer." *Scientific Reports* 11 (1): 10688.

Cornelissen, Germaine. 2014. "Cosinor-Based Rhythmometry." *Theoretical Biology & Medical Modelling* 11 (1): 16.

Ferguson, Amy, Laura M. Lyall, Joey Ward, et al. 2018. "Genome-Wide Association Study of Circadian Rhythmicity in 71,500 UK Biobank Participants and Polygenic Association with Mood Instability." *EBioMedicine* 35 (September): 279–287.

Finucane, Hilary K., Brendan Bulik-Sullivan, Alexander Gusev, et al. 2015. "Partitioning Heritability by Functional Annotation Using Genome-Wide Association Summary Statistics." *Nature Genetics* 47 (11): 1228–1235.

Friedman, Jerome, Trevor Hastie, and Rob Tibshirani. 2010. "Regularization Paths for Generalized Linear Models via Coordinate Descent." *Journal of Statistical Software* 33 (1): 1–22.

GTEx Consortium. 2020. "The GTEx Consortium Atlas of Genetic Regulatory Effects across Human Tissues." *Science (New York, N.Y.)* 369 (6509): 1318–1330.

Hemani, Gibran, Jie Zheng, Benjamin Elsworth, et al. 2018. "The MR-Base Platform Supports Systematic Causal Inference across the Human Phenome." *eLife* 7 (May). https://doi.org/10.7554/eLife.34408.

Honkanen, Jarno, Veera A. Timonen, Jessica R. Koski, et al. 2025. "Blood Donor Biobank Pipeline to Collect Genome-Based Samples for Research." In *Molecular Biology*, No. Biorxiv;2025.09.30.679473v1. BioRxiv, October 2. https://www.biorxiv.org/content/10.1101/2025.09.30.679473v1.full.

Ishigaki, Kazuyoshi, Saori Sakaue, Chikashi Terao, et al. 2022. "Multi-Ancestry Genome-Wide Association Analyses Identify Novel Genetic Mechanisms in Rheumatoid Arthritis." *Nature Genetics* 54 (11): 1640–1651.

Jansen, Iris E., Jeanne E. Savage, Kyoko Watanabe, et al. 2019. "Genome-Wide Meta-Analysis Identifies New Loci and Functional Pathways Influencing Alzheimer's Disease Risk." *Nature Genetics* 51 (3): 404–413.

Jansen, Philip R., Kyoko Watanabe, Sven Stringer, et al. 2019. "Genome-Wide Analysis of Insomnia in 1,331,010 Individuals Identifies New Risk Loci and Functional Pathways." *Nature Genetics* 51 (3): 394–403.

Jiang, Longda, Zhili Zheng, Ting Qi, et al. 2019. "A Resource-Efficient Tool for Mixed Model Association Analysis of Large-Scale Data." *Nature Genetics* 51 (12): 1749–1755.

Jones, Samuel E., Vincent T. van Hees, Diego R. Mazzotti, et al. 2019. "Genetic Studies of Accelerometer-Based Sleep Measures Yield New Insights into Human Sleep Behaviour." *Nature Communications* 10 (1): 1585.

Jones, Samuel E., Jacqueline M. Lane, Andrew R. Wood, et al. 2019. "Genome-Wide Association Analyses of Chronotype in 697,828 Individuals Provides Insights into Circadian Rhythms." *Nature Communications* 10 (1): 343.

Julkunen, Heli, Anna Cichońska, Mika Tiainen, et al. 2023. "Atlas of Plasma NMR Biomarkers for Health and Disease in 118,461 Individuals from the UK Biobank." *Nature Communications* 14 (1): 604.

Karczewski, Konrad J., Rahul Gupta, Masahiro Kanai, et al. 2024. "Pan-UK Biobank GWAS Improves Discovery, Analysis of Genetic Architecture, and Resolution into Ancestry-Enriched Effects." In *Genetic and Genomic Medicine*, No. Medrxiv;2024.03.13.24303864v2. MedRxiv, March 15. https://www.medrxiv.org/content/10.1101/2024.03.13.24303864v2.

Ke, Guolin, Qi Meng, Thomas Finley, et al. 2017. "LightGBM: A Highly Efficient Gradient Boosting Decision Tree." *Advances in Neural Information Processing Systems* 30. https://papers.nips.cc/paper_files/paper/2017/hash/6449f44a102fde848669bdd9eb6b76fa-Abstract.html.

Kyle, Simon D., Claire E. Sexton, Bernd Feige, et al. 2017. "Sleep and Cognitive Performance: Cross-Sectional Associations in the UK Biobank." *Sleep Medicine* 38 (October): 85–91.

Lagou, Vasiliki, Longda Jiang, Anna Ulrich, et al. 2023. "GWAS of Random Glucose in 476,326 Individuals Provide Insights into Diabetes Pathophysiology, Complications and Treatment Stratification." *Nature Genetics* 55 (9): 1448–1461.

Locke, Adam E., Bratati Kahali, Sonja I. Berndt, et al. 2015. "Genetic Studies of Body Mass Index Yield New Insights for Obesity Biology." *Nature* 518 (7538): 197–206.

Longchamps, R. J., S. Y. Yang, C. A. Castellani, et al. 2022. "Genome-Wide Analysis of Mitochondrial DNA Copy Number Reveals Loci Implicated in Nucleotide Metabolism, Platelet Activation, and Megakaryocyte Proliferation." *Human Genetics* 141 (1): 127–146.

Major Depressive Disorder Working Group of the Psychiatric Genomics Consortium. Electronic address:, and Major Depressive Disorder Working Group of the Psychiatric Genomics Consortium. 2025. "Trans-Ancestry Genome-Wide Study of Depression Identifies 697 Associations Implicating Cell Types and Pharmacotherapies." *Cell* 188 (3): 640–652.e9.

O'Connell, Kevin S., Maria Koromina, Tracey van der Veen, et al. 2025. "Genomics Yields Biological and Phenotypic Insights into Bipolar Disorder." *Nature* 639 (8056): 968–975.

Sinnott-Armstrong, Nasa, Yosuke Tanigawa, David Amar, et al. 2021. "Genetics of 35 Blood and Urine Biomarkers in the UK Biobank." *Nature Genetics* 53 (2): 185–194.

Specht, Adrien, German Kolosov, Katie L. J. Cederberg, et al. 2024. "Circadian Protein Expression Patterns in Healthy Young Adults." *Sleep Health* 10 (1S): S41–S51.

Sun, Benjamin B., Joshua Chiou, Matthew Traylor, et al. 2023. "Plasma Proteomic Associations with Genetics and Health in the UK Biobank." *Nature* 622 (7982): 329–338.

Surendran, Praveen, Elena V. Feofanova, Najim Lahrouchi, et al. 2020. "Discovery of Rare Variants Associated with Blood Pressure Regulation through Meta-Analysis of 1.3 Million Individuals." *Nature Genetics* 52 (12): 1314–1332.

Suzuki, Ken, Konstantinos Hatzikotoulas, Lorraine Southam, et al. 2024. "Genetic Drivers of Heterogeneity in Type 2 Diabetes Pathophysiology." *Nature* 627 (8003): 347–357.

Trubetskoy, Vassily, Antonio F. Pardiñas, Ting Qi, et al. 2022. "Mapping Genomic Loci Implicates Genes and Synaptic Biology in Schizophrenia." *Nature* 604 (7906): 502–508.

Weissbrod, Omer, Farhad Hormozdiari, Christian Benner, et al. 2020. "Functionally Informed Fine-Mapping and Polygenic Localization of Complex Trait Heritability." *Nature Genetics* 52 (12): 1355–1363.

Wu, Yeda, Enda M. Byrne, Zhili Zheng, et al. 2019. "Genome-Wide Association Study of Medication-Use and Associated Disease in the UK Biobank." *Nature Communications* 10 (1): 1891.

Yang, Jian, Beben Benyamin, Brian P. McEvoy, et al. 2010. "Common SNPs Explain a Large Proportion of the Heritability for Human Height." *Nature Genetics* 42 (7): 565–569.

Yang, Jian, Teresa Ferreira, Andrew P. Morris, et al. 2012. "Conditional and Joint Multiple-SNP Analysis of GWAS Summary Statistics Identifies Additional Variants Influencing Complex Traits." *Nature Genetics* 44 (4): 369–75, S1–3.

Yang, Jian, S. Hong Lee, Michael E. Goddard, and Peter M. Visscher. 2011. "GCTA: A Tool for Genome-Wide Complex Trait Analysis." In *The American Journal of Human Genetics*, vol. 88, 88. no. 1. Preprint. https://doi.org/10.1016/j.ajhg.2010.11.011.

Zheng, Zhili, Shouye Liu, Julia Sidorenko, et al. 2022. "Leveraging Functional Genomic Annotations and

Genome Coverage to Improve Polygenic Prediction of Complex Traits within and between Ancestries." In *bioRxiv*. October 14. https://doi.org/10.1101/2022.10.12.510418.
